# Vigorous intermittent lifestyle physical activity (VILPA) and mortality risk among US adults: a wearables-based national cohort study

**DOI:** 10.1101/2025.08.05.25333017

**Authors:** Nicholas A. Koemel, Matthew N. Ahmadi, Raaj Kishore Biswas, Cecilie Thøgersen-Ntoumani, Armando Texeira-Pinto, Clara K Chow, Jaroslaw Harezlak, Emmanuel Stamatakis

**Affiliations:** Mackenzie Wearables Research Hub, Charles Perkins Centre, The University of Sydney, Sydney, New South Wales, Australia; School of Health Sciences, Faculty of Medicine and Health, The University of Sydney, Sydney, New South Wales, Australia; Charles Perkins Centre, The University of Sydney, Sydney, New South Wales, Australia; Danish Centre for Motivation and Behaviour Science (DRIVEN), Department of Sports Science and Clinical Biomechanics, University of Southern Denmark, Odense, Denmark; School of Public Health, Faculty of Medicine and Health, University of Sydney, Australia; Westmead Applied Research Centre, University of Sydney and Department of Cardiology, Westmead Hospital, Sydney, Australia; Department of Epidemiology and Biostatistics, School of Public Health, Indiana University, Bloomington, IN, United States of America

**Author notes:** Corresponding author: Emmanuel Stamatakis, Address for correspondence: Hub D17, Charles Perkins Centre L6 West, the University of Sydney, New South Wales, Australia.

**Keywords:** Lifestyle physical activity, all-cause mortality, machine learning

## Abstract

**Background:** Vigorous intermittent lifestyle physical activity (VILPA) completed through normal daily living may offer a time-efficient avenue to accrue physical activity in a behaviorally sustainable manner. However, no research to date has explored its association with mortality in a nationally representative population. This study aimed to examine the dose-response association between VILPA and mortality risk in a nationally representative sample of US adults.

**Methods:** This study included a nationally representative sample of 3,293 US adults from the 2011-14 National Health and Nutrition Examination Survey (NHANES) who self-reported no participation in structured exercise (52.3% female; mean age: 50.7 [SD: 16.6 years]. The dose-response relationship between VILPA and all-cause mortality was estimated using multivariable-adjusted cubic splines. Average daily frequency (bouts/day) and duration (minutes/day) of VILPA bouts lasting up to one minute were measured using a wrist-worn accelerometer.

**Results:** Over the mean (SD) 6.7 (1.4) year follow-up period, 290 all-cause mortality events occurred. Compared to the referent point (0 bouts per day), there was an L-shaped dose-response association where the median frequency (5.3 bouts per day) was associated with a 44% lower risk of all-cause mortality (HR: 0.56; 95% CI: 0.39, 0.82). The dose response curve was less steep beyond approximately 8 bouts per day (HR: 0.46; 95% CI: 0.28, 0.77). Findings for the median frequency of VILPA bouts (5.3 bouts per day) remained consistent after excluding participants with poor health (HR: 0.51: 95% CI: 0.29, 0.87) and those who completed no VILPA (HR: 0.57: 95% CI: 0.38, 0.86). When excluding adults with prevalent cardiovascular disease or cancer at baseline (n= 2,731, 152 events), the dose-response relationship was similar, although the 95% CIs crossed unity for most of the curve (e.g. median frequency of VILPA bouts HR: 0.68: 95% CI: 0.41, 1.11).

**Conclusions:** In a nationally representative sample of US adults, short bursts of intermittent vigorous physical activity were associated with a lower risk of mortality. While these results highlight the potential of VILPA as a time-efficient source of activity, additional observational studies with longer follow up and larger sample sizes are warranted.

## INTRODUCTION

Physical inactivity is a major risk factor for all-cause, cardiovascular, and cancer mortality^1^, presenting an unresolved public health challenge. Despite decades of public health efforts, only approximately 20-25% of adults engage regularly in leisure time physical activity^2,3^. Both previous^4,5^ and current iterations^6,7^ of physical activity guidelines are largely derived from self-report questionnaires, primarily capturing activities lasting >10 minutes. Recent guidelines such as the 2018 Physical Activity Guidelines for Americans^6^ and the World Health Organization 2020 Guidelines on Physical Activity and Sedentary Behavior^7^, promote “all activity counts” messaging, implicitly recommending short bouts of physical activity. These recommendations were primarily informed by cross-sectional studies that generically assessed non-contextual and non-intensity-specific bouts lasting <10 minutes^8^, with no direct evidence of the role of physical activity coming from very short bouts.

Recent wearable based evidence from a sample of non-exercising adults in the UK Biobank has highlighted the potential health value of such brief bursts, specifically in the form vigorous intermittent lifestyle physical activity (VILPA: bouts lasting up to 1 minute each) and moderate-to-vigorous intermittent lifestyle physical activity (MV-ILPA: bouts <3 minutes)^9–12^. For example, 4.4 minutes per day of VILPA was associated with a 26-30% lower risk of all-cause and cancer mortality and a 32-34% lower risk of cardiovascular mortality^9^. Such studies incorporating wearable devices and novel machine learning methods^9–13^ permit the study of high-resolution “micro-patterns” of physical activity^9,10^, such as VILPA and MV-ILPA outlined above. This more granular examination of physical activity patterns was not previously possible in studies relying on self-report-based measures or even previous generation accelerometry studies^14^ with lower data resolution (1 min epoch).

From a behavioral standpoint, intermittent short bursts of incidental physical activity offer a theoretical advantage as they can be easily integrated into day-to-day life by individuals who face common barriers to traditional exercise such as a lack of time, motivation, or access to facilities^15^. Previous studies^9,11–13^ exploring the relationship between VILPA and mortality used the UK Biobank, a cohort that is more educated, has fewer chronic health conditions, less obesity, and is more socioeconomically advantaged compared to the general population^16^. To date, no study has examined the association of brief intermittent bursts of vigorous physical activity incorporated into daily living activities with mortality among a nationally representative dataset of US adults.

The aim of this study is to examine the dose-response association between daily VILPA frequency and duration with all-cause mortality risk in a nationally representative sample of US adults.

## METHODS

### Study Design and Participants

This prospective cohort included non-exercising adults aged ≥20 from the National Health and Nutrition Examination Survey (NHANES) during 2011-2014. NHANES is an ongoing annually collected survey that employs a multi-stage probability sampling design to capture the health and lifestyle habits representative of the noninstitutionalized US population^17^. This sampling design includes oversampling certain subgroups, such as racial/ethnic minorities, low-income individuals, and older adults, to improve the precision of prevalence estimates in these subgroups when incorporating survey weights that account for oversampling, non-response and poststratification^17^. All NHANES protocols have been approved by the National Center for Health Statistics and informed consent was collected from each participant.

From 2011-2014, during the mobile examination visits, a sub-sample of 14,693 participants from the main NHANES survey were invited to wear wrist-worn trackers (ActiGraph model GT3X+, Pensacola, FL) on their non-dominant hand for 7-days^17–19^. Participants were instructed to wear the devices for 24 hours per day while sleeping and awake, including while in the bath, shower, or when swimming. The devices were programmed with a sampling rate of 80 Hz and distributed to participants at the end of the mobile examination visits^17–19^. To be included in our analyses, participants must have worn the device at least three valid monitoring days (>16 hours), including at least one weekend day^9–12,20,21^. We excluded all subjects who incorrectly wore the device on the dominant wrist, reported mobility issues such as difficulty walking, or required equipment to walk (**Supplement Fig. 1**)^22^.

As in previous studies^9–13,23,24^, to enable the examination of the health effects of incidental physical activity, we included only participants who reported no moderate or vigorous leisure time exercise participation in a typical week, as captured by the Global Physical Activity Questionnaire at baseline^17–19^. The examples provided include participating in recreational sport activities such as running, basketball, golf, bicycling, or walking (**Supplement Table 1**).

### Ascertainment of Mortality

Mortality data was ascertained via the National Center for Health Statistics where participant information was linked to the National Death Index. The underlying cause of death was reported using the International Statistical Classification of Diseases and Related Health Problems (ICD-10) code. All-cause mortality included deaths from all known causes (**Supplement Table 2**). The follow-up period was defined as the duration from the mobile examination (i.e., date of accelerometry collection) to the date of death or December 31^st^, 2019 for those without a mortality event. Analyses were performed between January 1^st^ and August 1^st^ 2024.

### Device-Measured Physical Activity Classification

As described in detail previously^9–13,25^, we used a validated two-stage random forest intensity classification schema to determine specific activities and intensity (84.6% accuracy across all intensities; see **Supplement Methods**). In brief^9–13,25^, this classifier first categorizes each 10-second window as one of four activity classes including: sedentary (lying or sitting still), standing utilitarian (for example, ironing a shirt, washing dishes), walking (for example, gardening, active commuting, mopping floors), or running/high energetic activities (for example, active playing with children). At the second stage, these activity classes are then assigned one of four intensities including: sedentary, light (<3 METs), moderate (≥3 to <6 METs), and vigorous activities (≥6 METs). Walking activities were classified using the raw magnitude of wrist movement acceleration using milligravity (mg), defined as light (<100 mg), moderate (≥100 mg) and vigorous (≥400 mg) intensity. The vigorous intensity definition and bout length selection of up to 1 minute in duration was based on both laboratory data and the naturally occurring bout length in the present and previous studies^9,11–13^. Additional details regarding the VILPA definition and bout length selection are provided in the **Supplemental Methods**.

### Statistical Analysis

The frequency and daily duration for VILPA were truncated at the 2.5 and 97.5 percentile to reduce the potential influence of sparse data^9,20^. We estimated multivariable adjusted absolute risk for all models using log-link Poisson Regression with robust standard error to calculate the dose-response association of VILPA frequency and daily duration with mortality risk^13,20^. We assessed the dose-response association between VILPA with mortality risk using restricted cubic splines with 0 bouts per day and 0 minutes per day for daily duration as the reference (knots placed at the 10^th^, 50^th^, and 90^th^ percentile)^9,10^. Time-to-event associations of VILPA with mortality risk were estimated using Cox proportional hazards models. To provide point estimates for the dose-response associations, we report the hazard ratio (HR) for the median of each exposure and the minimal volume dose (50% of the optimal dose)^9,20,26^. We identified the inflection point^27^ in the dose-response curve by examining the difference between successive HRs (**Supplement Fig. 2-3**). To account for the complex survey design, we applied the weights to all models which ensures the estimates are representative of the noninstitutionalized US population^17^. All models met the Cox proportional hazards assumptions.

As in previous analogous analyses^9,10,13,20^, we adjusted for self-reported age, gender, education, race/ethnicity, socioeconomic status (income-to-poverty ratio), fruit and vegetable intake (servings per day), accelerometry estimated sleep duration, medication (glycemic control, lipid-lowering, or blood pressure), discretionary screen time, previous cancer, previous cardiovascular disease, previous diabetes, and familial history of cardiovascular disease or type-two diabetes^9,10,13,20^. A detailed description of each covariate is provided in **Supplement Table 3**. The models with VILPA frequency (bouts per day) as the exposure were additionally adjusted for total physical activity energy expenditure (PAEE)^28,29^. The models with VILPA duration (minutes per day) as the exposure were additionally adjusted for light and moderate PAEE and the daily VILPA duration from bout lengths >1 minute^9–11,13^. All missing covariates were estimated using a random forest multiple imputation by chained equations model (5 imputed datasets).

### Sensitivity Analyses

To test for the possibility of sex-based differences, we tested for multiplicative interaction and the relative excess risk due to interaction. We completed several sensitivity analyses to examine the robustness of our findings to reverse causation and other possible biases. First, we excluded underweight participants (BMI of <18.5), participants reporting a self-perceived rating of poor health or participants with prevalent CVD or cancer. Second, we created additional models using the median value of the exposure as the reference point in the dose-response association^10,20^ or excluding individuals with extreme VILPA values. Third, we repeated the main analyses with adjustment for cardiometabolic risk factors, which are possible mediators in the association between physical activity and mortality risk^10,30^. We provide a sensitivity analysis adjusting for a more comprehensive diet quality indicator (Healthy Eating Index 2015)^31^ to address potential confounding by other dietary factors^22^. In another analysis, we excluded individuals with missing covariate information rather than applying multiple imputations to address potential imputation bias^32^. To test for possible misclassification of non-exercisers, we completed a sensitivity analysis repeating the main analyses with two alternative non-exerciser definitions a) excluding adults who self-reported only completing recreational VPA and b) excluding adults who self-reported ≥1 day per week of recreational MVPA). We also conducted a sensitivity analysis by treating accidents and residual deaths (i.e., contains a mixture of accidental and other causes of deaths that do not have clear links to physical inactivity) as a competing interest using Fine-Gray sub-distribution hazard models (see **Supplement Table 2** for ICD-10 Codes).

### Additional Analyses

To enable a direct comparison of VILPA associations with our previous work in middle-aged adults^9–13^, we repeated the core analysis in adults ≥40 years old. Additionally, to assess the possibility of unmeasured confounding, we calculated e-values for each model^33^. There was no problematic multicollinearity present in any of the models (**Supplement Table 4**).

All analyses and graphics in this study were conducted using the survival, survey, RMS, mice, and ggplot2 packages in R statistical software (v.4.3.1). This study followed the Strengthening the Reporting of Observational Studies in Epidemiology Guidelines (STROBE).

## RESULTS

### Participant characteristics

The final analytical sample included 3,293 non-exercising adults (52.3% female) with a median age of 50.7 [SD: 16.6] years (**Table 1**). The sample included a substantial proportion of adults (44%) consuming medication for glycemic control, lipid lowering, or blood pressure management, with 9.8% reporting a history of cardiovascular disease and 9.3% reporting a history of cancer. The majority of participants had complete covariate information (92.9%). The vast majority of all vigorous bouts lasted <1 minute in duration (95% of all bouts) with a median bout length of 10 seconds [IQR: 20 seconds]. The median VILPA frequency and duration were 5.3 [IQR: 8.9] bouts per day and 1.1 minutes [IQR: 2.09] per day, respectively. Participants had a median wear time of 23.1 [IQR: 1.2] hours per day. Over a 6.7-year median follow-up period, there were 290 all-cause mortality events, corresponding to 22,183 person-years.

**Table 1:** Participant baseline characteristics by VILPA duration (mins/day).

| **VILPA duration quartiles**<br>**(mins/day)** | **0** | **<0.4** | **0.4-1.1** | **1.1-2.5** | **>2.5** | **Overall** |
| --- | --- | --- | --- | --- | --- | --- |
| **Sample size (n)** | 129 | 791 | 791 | 791 | 791 | 3,293 |
| **All-cause mortality, n (%)** | 59 (45.7) | 129 (16.3) | 60 (7.6) | 26 (3.3) | 16 (2.0) | 290 (8.8) |
| **Follow up (in years), mean**<br>**(SD)** | 5.8 (1.8) | 6.5 (1.6) | 6.8 (1.4) | 6.8 (1.3) | 6.9 (1.2) | 6.7 (1.4) |
| **Age (mean (SD))** | 72.6 (9.2) | 62.4 (14.3) | 49.9 (14.9) | 45.2 (14.4) | 41.8 (13.2) | 50.7 (16.6) |
| **Sex, Female (%)** | 68 (52.7) | 468 (59.2) | 455 (57.5) | 404 (51.1) | 327 (41.3) | 1,722 (52.3) |
| **Smoking (%)** |  |  |  |  |  |  |
| Current | 24 (18.6) | 167 (21.1) | 235 (29.7) | 187 (23.7) | 190 (24.1) | 803 (24.4) |
| Never | 61 (47.3) | 397 (50.3) | 399 (50.4) | 451 (57.2) | 451 (57.1) | 1,759 (53.5) |
| Previous | 44 (34.1) | 226 (28.6) | 157 (19.8) | 151 (19.1) | 149 (18.9) | 727 (22.1) |
| **Alcohol, No (%)<sup>1</sup>** | 53 (41.1) | 279 (35.3) | 223 (28.2) | 230 (29.1) | 203 (25.7) | 988 (30.0) |

| **Education (%)** |  |  |  |  |  |  |
| --- | --- | --- | --- | --- | --- | --- |
| Less than 9th grade | 21 (16.3) | 105 (13.3) | 64 (8.1) | 72 (9.1) | 116 (14.7) | 378 (11.5) |
| 9-11th grade | 30 (23.3) | 135 (17.1) | 119 (15.0) | 138 (17.4) | 154 (19.5) | 576 (17.5) |
| High school graduate/GED or equivalent | 28 (21.7) | 179 (22.6) | 194 (24.5) | 177 (22.4) | 194 (24.5) | 772 (23.4) |
| Some college or AA degree | 34 (26.4) | 232 (29.3) | 267 (33.8) | 242 (30.6) | 217 (27.4) | 992 (30.1) |
| College graduate or above | 16 (12.4) | 140 (17.7) | 147 (18.6) | 162 (20.5) | 110 (13.9) | 575 (17.5) |
| **Ethnicity (%)** |  |  |  |  |  |  |
| Mexican American | 5 (3.9) | 66 (8.3) | 96 (12.1) | 119 (15.0) | 172 (21.7) | 458 (13.9) |
| Hispanic | 10 (7.8) | 71 (9.0) | 91 (11.5) | 98 (12.4) | 101 (12.8) | 371 (11.3) |
| White | 69 (53.5) | 392 (49.6) | 322 (40.7) | 272 (34.4) | 228 (28.8) | 1,283 (39.0) |
| Black | 39 (30.2) | 203 (25.7) | 185 (23.4) | 167 (21.1) | 168 (21.2) | 762 (23.1) |
| Asian | 5 (3.9) | 50 (6.3) | 74 (9.4) | 108 (13.7) | 108 (13.7) | 345 (10.5) |
| Multi-Racial | 1 (0.8) | 9 (1.1) | 23 (2.9) | 27 (3.4) | 14 (1.8) | 74 (2.2) |
| **Diet<sup>2</sup>** | 4.3 (2.8) | 4.2 (2.9) | 4.3 (3.1) | 4.3 (2.8) | 4.4 (3.6) | 4.3 (3.1) |
| **Income to poverty ratio (mean (SD))** | 2.2 (1.4) | 2.4 (1.6) | 2.2 (1.6) | 2.3 (1.6) | 2.1 (1.5) | 2.2 (1.6) |
| **Medication for cholesterol, glycemic control and blood pressure, Yes (%)** | 113 (87.6) | 529 (66.9) | 358 (45.3) | 268 (33.9) | 181 (22.9) | 1,449 (44.0) |
| **No previous CVD (%)** | 72 (55.8) | 645 (81.5) | 741 (93.7) | 750 (94.8) | 763 (96.5) | 2,971 (90.2) |
| **No previous Cancer (%)** | 91 (70.5) | 668 (84.5) | 726 (91.8) | 743 (93.9) | 760 (96.1) | 2,988 (90.7) |
| **No familial CVD** | 111 (86.0) | 694 (87.7) | 686 (86.7) | 719 (90.9) | 719 (90.9) | 2,929 (88.9) |
| **No familial diabetes** | 80 (62.0) | 431 (54.5) | 433 (54.7) | 504 (63.7) | 483 (61.1) | 1,931 (58.6) |
| **LPA (mean (SD))** | 46.0 (17.5) | 59.5 (21.0) | 66.6 (21.1) | 71.5 (20.9) | 74.6 (21.2) | 67.2 (22.0) |
| **MPA (mean (SD))** | 1.8 (3.9) | 6.0 (7.5) | 13.1 (10.7) | 19.0 (12.6) | 29.4 (16.7) | 16.3 (15.0) |
| **Sleep (mean (SD))** | 359.2 (134.9) | 365.0 (127.1) | 386.2 (113.8) | 388.1 (110.9) | 391.0 (104.9) | 381.7 (115.8) |
| **BMI (mean (SD))** | 29.4 (6.6) | 30.2 (6.9) | 30.0 (6.9) | 29.1 (6.5) | 28.1 (5.9) | 29.4 (6.6) |

VILPA: Vigorous intermittent lifestyle physical activity; MPA: Moderate intensity physical activity; LPA: light intensity physical activity.
The columns breakdown corresponds to the total volume of VILPA. Values represent mean (standard deviation) unless specified otherwise. <sup>1</sup>Alcohol consumption measured 12 drinks/year (1 drink = 12 oz. beer/ 5 oz. glass of wine/ one and a half oz. of liquor). <sup>2</sup>Fruits and vegetable consumption measured in servings per day.

### VILPA and all-cause mortality risk

In the fully adjusted analytical model, both VILPA frequency and duration displayed an L-shaped association with lower all-cause mortality risk. The minimum daily VILPA frequency (0 bouts per day, 9.6% of the sample) and median (5.3 bouts per day) frequency of VILPA were associated with an absolute risk of 95.7 (95% CI: 55.3, 165.4) and 55.0 (95% CI: 33.5, 90.2) per 10,000 person-years; respectively (**Supplement Fig. 4A**). The minimum daily VILPA duration (0 minutes per day; 25.1% of the sample) was associated with an absolute risk of 89.8 (95% CI: 52.5, 153.4) per 10,000 person-years. The median daily VILPA duration (1.1 minutes per day) had an absolute risk of 55.7 (95% CI: 34.0, 91.3; **Supplement Fig. 4B**) per 10,000 person-years.

Compared to the VILPA frequency referent data point (0 bouts per day), we observed an L-shaped dose-response association between VILPA frequency and all-cause mortality risk (**Fig. 1A**). The median daily VILPA frequency (5.3 bouts per day) was associated with an HR of 0.56 (95% CI: 0.39, 0.82) for all-cause mortality. The slope of the dose-response curve for VILPA bouts per day began to plateau beyond approximately 8 bouts per day, corresponding to a 54% lower risk of all-cause mortality (HR: 0.46; 95% CI: 0.28, 0.77; **Supplement Fig. 2**). Compared to the referent minimum daily VILPA duration (0 minutes per day), there was a near-linear association with all-cause mortality risk (**Fig. 1B**). The median daily duration of VILPA (1.1 minutes/day) was associated with an HR of 0.61 (95% CI: 0.42, 0.88). The slope of the dose-response relationship began to plateau beyond approximately 2 minutes per day, corresponding to an HR of 0.55 (95%CI: 0.27, 0.76; **Supplement Fig. 3**). The minimal dose for VILPA bouts (4.3 bouts per day) and daily duration (1.3 minutes per day) corresponded to an HR of 0.62 (95%CI: 0.44, 0.86) and 0.57 (95%CI: 0.37, 0.85), respectively.

**Figure 1:**
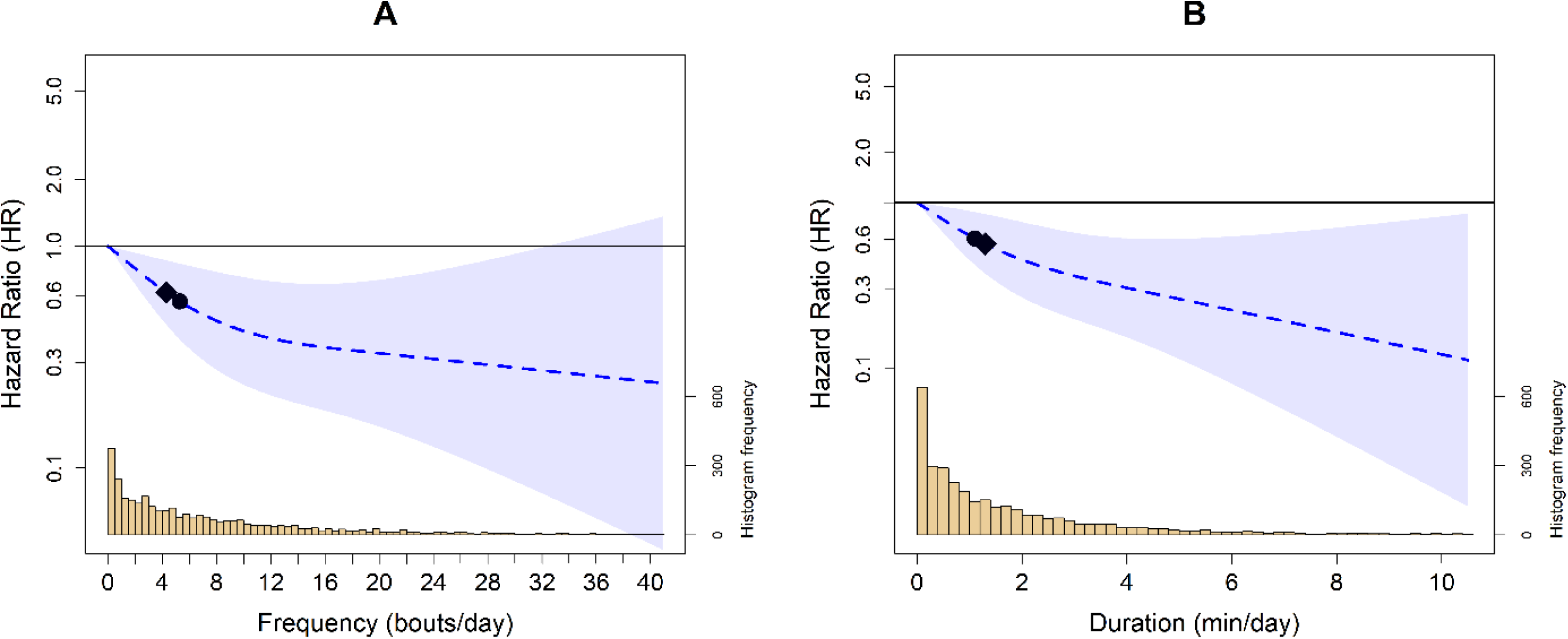
Survey adjusted dose response curves of daily VILPA frequency (A) and duration (B) with all-cause mortality (n = 3,293; events = 290). **Legend**: Analyses were adjusted for sex, age, income, education, ethnicity, fruit and vegetable consumption, smoking hist ory, physical activity energy expenditure from LPA and MPA, alcohol consumption, sleep duration, duration of light and mo derate intensity, discretionary screentime, medication use (glycemic control, blood pressure, cholesterol), family history of diabetes and CVD, and previous history of CVD, diabetes, and cancer. VILPA bout (A) was further adjusted by energy exp enditure by vigorous intensity and VILPA duration (B) was further adjusted for VILPA bouts over 1-minute bout. All analyse s excluded participants who had an event in the first year of follow-up. All analyses were adjusted for strata, cluster, and su rvey weights. Reference was set to zero for VILPA frequency and duration. Diamond, minimal dose, as indicated by the ED 50 statistic which estimates the daily duration of VILPA associated with 50% of optimal risk reduction. Circle, HR associate d with the median VILPA value

In adults ≥40 years old, the association between VILPA frequency and duration was attenuated, displaying a clearer L-shape relationship with all-cause mortality (**Fig. 2**). Compared to the minimum VILPA frequency (0 bouts per day) and duration (0 minutes per day), the median VILPA frequency (3.8 bouts per day) and duration (0.8 minutes per day) were associated with an HR of 0.61 (95%CI: 0.43, 0.86) and 0.65 (95%CI: 0.47, 0.90), respectively. The minimal dose for VILPA frequency (3.2 bouts per day) and daily duration (1.1 minutes per day) was associated with an HR of 0.65 (95%CI: 0.48, 0.88) and 0.58 (95%CI: 0.39, 0.87), respectively.

**Figure 2:**
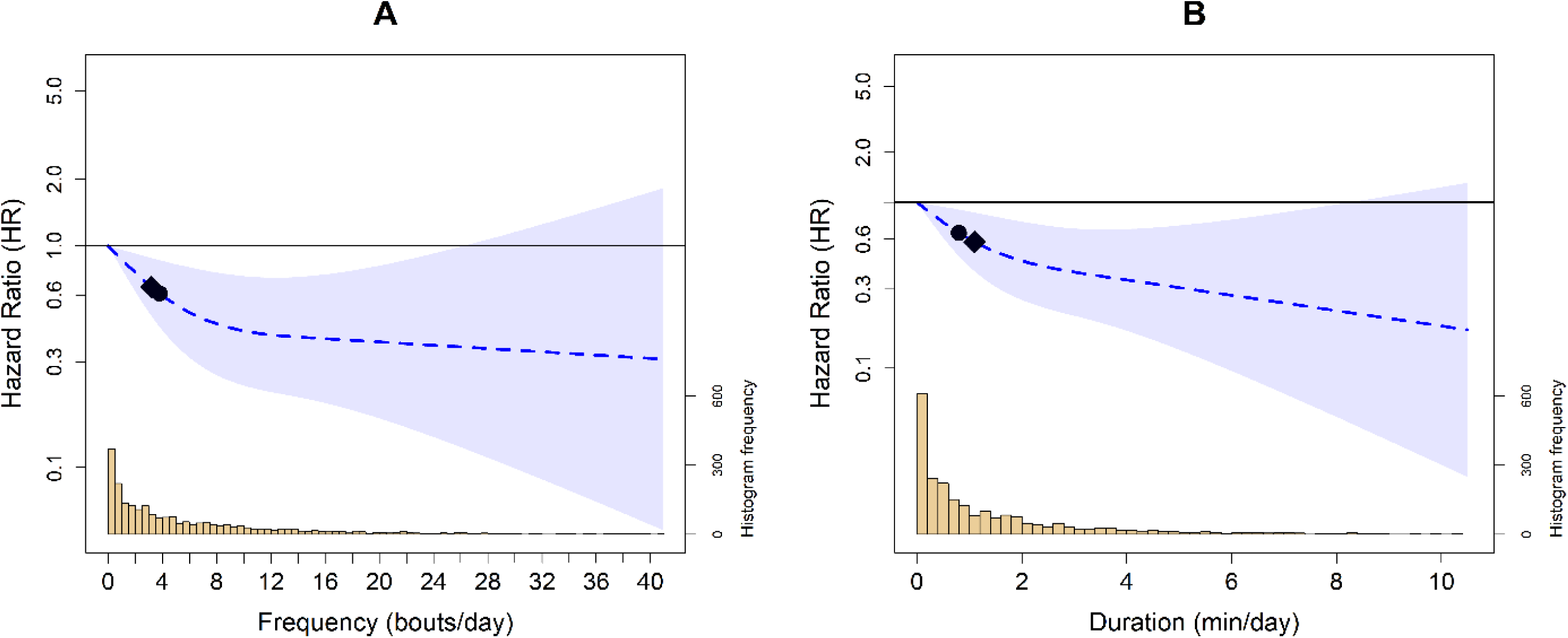
Survey adjusted dose response curves of daily VILPA frequency (A) and duration (B) with all-cause mortality (n = 2,276; events = 272) in adults aged ≥40. **Legend**: Analyses were adjusted for sex, age, income, education, ethnicity, fruit and vegetable consumption, smoking history, physical activity energy expenditure from LPA and MPA, VILPA bouts over 1-minute, alcohol consumption, sleep duration, duration of light and moderate intensity, discretionary screentime, medication use (glycemic control, blood pressure, cholesterol), family history of diabetes and CVD, and previous history of CVD, diabetes, and cancer. VILPA bout (A) was further adjusted by energy expenditure by vigorous intensity and VILPA duration (B) was further adjusted for VILPA bouts over 1-minute bout. Reference was set to zero for VILPA frequency and duration. Diamond, minimal dose, as indicated by the ED50 statistic which estimates the daily duration of VILPA associated with 50% of optimal risk reduction. Circle, HR associated with the median VILPA value. All analyses excluded participants who had an event in the first year of follow-up. All analyses were adjusted for strata, cluster, and survey weights.

### Sensitivity analyses

The associations between VILPA and all-cause mortality did not significantly differ when excluding individuals with poor self-rated health (**Supplement Fig. 5**). When excluding adults with prevalent disease (**Supplement Fig. 6**), the dose-response curves were similar to main analyses but 95%CIs crossed unity for most of the curve. The associations were not materially affected when adjusting for cardiometabolic health (**Supplement Figs. 7-8**), diet quality (**Supplement Fig. 9**), or when excluding individuals with upper and lower ranges of VILPA (**Supplement Figs. 10-12**). The associations were unchanged when treating accidents as a competing interest, while treating residual deaths as a competing interest largely attenuated the associations (**Supplement Figs. 13-14**). There were no materially different results when using the median as the referent point (**Supplement Fig. 15**), different definitions of non-exercisers (**Supplement Figs. 16-17**), or when excluding individuals with missing covariate information (**Supplement Fig. 18**).

### Additional Analyses

There was no significant interactive or synergistic association between sex and VILPA with all-cause mortality (**Supplement Table 5**). E-values indicated that to attenuate the primary findings to null, the association of an unmeasured confounder for VILPA frequency and duration would need to be 3.59 (lower 95% CI: 1.96) and 3.59 (lower 95% CI: 2.00), respectively (**Supplement Table 6**).

## DISCUSSION

### Statement of Principal Findings

This is the first US-based study on the associations of brief intermittent vigorous activity embedded into daily living with mortality risk. We found that daily VILPA frequency was inversely associated with all-cause mortality such that accumulating 5-6 very brief bouts lasting <1 minute each, was associated with a 42-47% lower risk of all-cause mortality. We also found a near-linear inverse association between VILPA duration and all-cause mortality risk, indicating that relatively modest amounts of VILPA, such as the median duration of just over one minute, were associated with a 39% lower risk of all-cause mortality. Our findings were generally supported by a range of sensitivity analyses addressing potential biases, including reverse causation. Although dose response curve was not materially affected when participants with CVD and cancer were excluded, the 95% confidence intervals were widened and crossed unity, a finding that raises some uncertainty relating to the influence of reverse causation on these findings.

Previous physical activity research using the US representative NHANES data has primarily focused on context-agnostic total physical activity in the form of total daily steps^34,35^ or MVPA^14,36–40^ and limited exploration of bouts lasting <10 minutes in duration^14,41^. To date, only two studies have examined the association between short bouts of MVPA (i.e., 5 or 10 minutes in duration) with all-cause mortality using prospective data^14,41^. This relatively underexplored aspect of physical activity is important, as the vast majority of adults complete their daily moderate or vigorous physical activity in brief bouts lasting <3 minutes in duration (e.g. 88% of all MVPA bouts in the UK Biobank^10,42^, and 92% of bouts in MVPA NHANES). This US study is the first, to our knowledge, in a population representative sample, examining the dose-response associations of brief bouts of VILPA^9^ lasting <1 minute in duration with all-cause mortality.

In comparison to the previous analogous work examining VILPA frequency and duration in the UK Biobank^9,10,13^, our findings indicate a larger magnitude of risk reduction per exposure unit. For example, the median VILPA of 4.4 minutes per day in the UK Biobank was associated with a 38% lower risk of all-cause mortality^9^, an effect size that was elicited in NHANES from 1.1 minutes per day. Moreover, VILPA frequency demonstrated steep beneficial L-shaped association with all-cause mortality where the steepness of the dose-response curve began to diminish beyond 8 bouts per day, associated with a 54% lower risk of all-cause mortality. Of note, the VILPA bouts in NHANES occurred less frequently (Median: 5.3 [IRQ: 8.9] bouts per day) and were shorter (Median: 10 seconds [IQR: 20 seconds] each) when compared to the UK Biobank (Median [IRQ] 10.1 [12.8] bouts per day and lasting 25 [6] seconds each).

The noticeable difference between the observed effect size for VILPA in the NHANES dataset and the UK Biobank may be partially attributed to variations in sociodemographic and lifestyle characteristics between the cohorts and the different prevalence of vigorous physical activity^16,43^. For example, compared to the non-exercisers in UK Biobank participants, the current NHANES sample had a higher average BMI (29.4 vs. 27.6), contained over twice the proportion of current smokers (24.4% vs. 9.2%), and completed approximately half the total daily MVPA (18.4 min vs. 30.3 min/day)^9,10,13^. These differences in cohort profiles likely reflect a combination of the non-representative nature of the UK Biobank, which compared to the general UK population, is more socioeconomically advantaged, less obese, less likely to smoke or drink, and reported fewer self-reported health conditions^9,10,13,16^ and the high prevalence of insufficient physical activity in the US^43^. These collective differences may contribute to a lower fitness and functional capacity in the NHANES sample, potentially resulting in a greater relative benefit from vigorous physical activity. Additional evidence from the UK Biobank further supports this in the context of incidental physical activity, wherein inactive adults achieved up to two-fold lower mortality risk from intermittent physical activity when compared to adults meeting the physical activity guidelines^42^.

This study expands on previous findings^14,38,39^ among US adults, by demonstrating meaningfully lower risk associated with modest amounts of vigorous-intensity physical activity accrued through very short bouts. While the magnitude of risk reduction observed in our study is substantial, it may be biologically feasible, particularly given this study examines a population of highly inactive and overweight individuals. Previous research has shown greater cardiorespiratory benefits from brief bursts of high-intensity training (HIIT) in obese or inactive adults when compared to healthy weight active adults^44^. These benefits include various cardiometabolic risk factors that additively contribute to mortality risk, including improved cardiorespiratory fitness, enhanced endothelial function, body composition, reduced blood pressure, and improved glycemic control^44,45^. While no current recommendations exist for the frequency of vigorous bouts per day, our findings align well with previous evidence suggesting that as little as 3-6 bouts per day of high-intensity movement lasting for 6-20 seconds (e.g., stair climbing) can improve cardiometabolic profiles in both young and older adults^46–48^.

The present results highlight some promise for brief, intermittent activity, especially considering that the majority of US adults may not be keen, able, or willing to engage in regular structured exercise^49^. Previous qualitative work^15^, indicates that interventions integrating such brief activity bursts into everyday tasks (e.g., active commuting, household chores, or office work) without the need for dedicated time or equipment may have feasibility advantages over traditional exercise. Commonly reported VILPA activities in a free-living environment include very fast walking, incline walking, short running bursts, outdoor gardening, carrying heavy loads during household chores, or energetic playing with children^10^. If supported by additional research, identifying additional VILPA activities that are both widely accessible and practical could create opportunities to inform public health initiatives or interventions aimed at promoting physical activity in those unable or unwilling to complete traditional structured exercise.

### Strengths and Limitations

A key strength of this study is the incorporation of survey design weights into our analyses enabling population-representative estimates for non-institutionalized adults in the US. We also employ a novel machine-learning approach that classifies physical activity intensity in 10-second windows^25,50^. This is distinct from previous NHANES context-agnostic work which has predominantly examined accelerometry-based total amounts of PA accrued through 1-2 min epochs which are unable to capture very short bouts of VILPA^14,34–41^, or self-reported leisure time physical activity^51–54^. We conducted an extensive range of additional analyses to test the robustness of our results to the risk of reverse causation, confounding, and other biases, including the exclusion of prevalent diseases, frail or poor health individuals, and adjustments for cardiometabolic health markers^9,10,20,22,55^. We also calculated e-values to assess the possibility of unmeasured confounding^33^ indicating that for an unmeasured confounder to attenuate the primary findings to null, the association of an unmeasured confounder for VILPA frequency and duration would need to be 3.59 (lower 95% CI: 1.96) and 3.59 (lower 95% CI: 2.00). These results suggest that such a confounder would need to have a strong and unlikely association with both the exposure and outcome to fully account for the observed relationships.

Despite the measures undertaken, the possibility of reverse causation or unmeasured confounding remains. In particular, we demonstrate that when excluding adults with prevalent CVD and cancer at baseline the dose-response relationship was attenuated, likely due to compromised statistical power and the loss of 138 events (48% of all events), given the high prevalence of CVD and cancer in the cohort and modest follow-up period (6.7 years). Factors such as misreporting, temporal variation of lifestyle factors (e.g., alcohol intake, smoking, obesity), or the high prevalence of chronic diseases at baseline could have influenced the study findings. Another limitation of this study also includes the possibility of incomplete capturing of VILPA using wrist-worn wearables, e.g. the intensity of activities such as walking uphill or carrying objects (e.g., backpacks) may not be fully captured^25^, likely leading to an underestimation of true daily VILPA. Such underestimation could in turn bias estimates, leading to an overestimation of the effect sizes per VILPA time unit.

## CONCLUSION

VILPA frequency and duration were associated with lower all-cause mortality risk in an L-shaped and linear manner, respectively. While these findings highlight the potential of VILPA as a time-efficient and behaviorally sustainable form of activity, further replication is needed in diverse cohorts with long follow up.

## Declarations

### Ethics approval and consent to participate

All NHANES protocols have been approved by the National Center for Health Statistics and informed consent was collected from each participant.

## Consent for publication

All data has been publicly consented for by participants enrolled in NHANES.

## Availability of data and materials

The data that support the findings of this study are publicly available at https://wwwn.cdc.gov/nchs/nhanes/.

## Authors’ contributions

NAK, RKB, MNA: conceptualization, methodology, software, formal analysis, validation, visualization, writing – original draft. CTN, ATP, CKC, JH: conceptualization, methodology, writing – review and editing. ES: funding acquisition, project administration, resources, supervision, writing – review and writing. ES is the guarantor, is responsible for the overall content and accepts full responsibility for the work and/or the conduct of the study, had access to the data and controlled the decision to publish. All authors read and approved the final manuscript. The corresponding author attests that all listed authors meet authorship criteria and that no others meeting the criteria have been omitted.

## References

1. Lear SA, Hu W, Rangarajan S, et al. The effect of physical activity on mortality and cardiovascular disease in 130 000 people from 17 high-income, middle-income, and low-income countries: the PURE study. The Lancet. 2017;390(10113):2643–2654. doi:10.1016/S0140-6736(17)31634-3

2. Guthold R, Stevens GA, Riley LM, Bull FC. Worldwide trends in insufficient physical activity from 2001 to 2016: a pooled analysis of 358 population-based surveys with 1.9 million participants. The Lancet Global Health. 2018;6(10):e1077–e1086. doi:10.1016/S2214-109X(18)30357-7

3. Stamatakis E, Chaudhury M. Temporal trends in adults’ sports participation patterns in England between 1997 and 2006: the Health Survey for England. British Journal of Sports Medicine. 2008;42(11):901. doi:10.1136/bjsm.2008.048082

4. Organization WH. Global recommendations on physical activity for health. World Health Organization; 2010.

5. Health UDo, Services H. US Department of Health and Human Services 2008 physical activity guidelines for Americans. Hyattsville, MD: Author, Washington, DC. 2008;2008:1–40.

6. Piercy KL, Troiano RP, Ballard RM, et al. The Physical Activity Guidelines for Americans. JAMA. 2018;320(19):2020–2028. doi:10.1001/jama.2018.14854

7. Bull FC, Al-Ansari SS, Biddle S, et al. World Health Organization 2020 guidelines on physical activity and sedentary behaviour. British Journal of Sports Medicine. 2020;54(24):1451–1462.

8. Jakicic JM, Kraus WE, Powell KE, et al. Association between Bout Duration of Physical Activity and Health: Systematic Review. Med Sci Sports Exerc. Jun 2019;51(6):1213–1219. doi:10.1249/mss.0000000000001933

9. Stamatakis E, Ahmadi MN, Gill JMR, et al. Association of wearable device-measured vigorous intermittent lifestyle physical activity with mortality. Nature Medicine. 2022;28(12):2521–2529. doi:10.1038/s41591-022-02100-x

10. Ahmadi MN, Hamer M, Gill JM, et al. Brief bouts of device-measured intermittent lifestyle physical activity and its association with major adverse cardiovascular events and mortality in people who do not exercise: a prospective cohort study. The Lancet Public Health. 2023;8(10):e800–e810.

11. Koemel NA, Ahmadi MN, Biswas RK, et al. Can incidental physical activity offset the deleterious associations of sedentary behaviour with major adverse cardiovascular events? European Journal of Preventive Cardiology. 2025;doi:10.1093/eurjpc/zwae316

12. Stamatakis E, Ahmadi MN, Biswas RK, et al. Device-measured vigorous intermittent lifestyle physical activity and major adverse cardiovascular events: evidence of sex differences. British Journal of Sports Medicine. 2024;doi:10.1101/2023.10.23.23297430

13. Stamatakis E, Ahmadi MN, Friedenreich CM, et al. Vigorous Intermittent Lifestyle Physical Activity and Cancer Incidence Among Nonexercising Adults: The UK Biobank Accelerometry Study. JAMA Oncology. 2023;9(9):1255–1259. doi:10.1001/jamaoncol.2023.1830

14. Ekelund U, Tarp J, Steene-Johannessen J, et al. Dose-response associations between accelerometry measured physical activity and sedentary time and all cause mortality: systematic review and harmonised meta-analysis. BMJ. 2019;366:l4570. doi:10.1136/bmj.l4570

15. Thøgersen-Ntoumani C, Kritz M, Grunseit A, et al. Barriers and enablers of vigorous intermittent lifestyle physical activity (VILPA) in physically inactive adults: a focus group study. International Journal of Behavioral Nutrition and Physical Activity. 2023;20(1):78. doi:10.1186/s12966-023-01480-8

16. Fry A, Littlejohns TJ, Sudlow C, et al. Comparison of sociodemographic and health-related characteristics of UK Biobank participants with those of the general population. American journal of epidemiology. 2017;186(9):1026–1034.

17. National Center for Health Statistics. NHANES Survey Methods and Analytic Guidelines. 2024

18. National Center for Health Statistics. National Health and Nutrition Examination Survey (NHANES) 2013-2014 Procedures Manual. https://wwwn.cdc.gov/nchs/nhanes/continuousnhanes/manuals.aspx?BeginYear=2013

19. National Center for Health Statistics. National Health and Nutrition Examination Survey (NHANES) 2011-2012 Procedures Manual. https://wwwn.cdc.gov/nchs/nhanes/continuousnhanes/manuals.aspx?BeginYear=2011

20. Ahmadi MN, Clare PJ, Katzmarzyk PT, del Pozo Cruz B, Lee IM, Stamatakis E. Vigorous physical activity, incident heart disease, and cancer: how little is enough? European Heart Journal. 2022;43(46):4801–4814. doi:10.1093/eurheartj/ehac572

21. Stamatakis E, Koemel NA, Biswas RK, et al. Minimum and optimal combined variations in sleep, physical activity, and nutrition in relation to all-cause mortality risk. BMC Medicine. 2025;23(1):111. doi:10.1186/s12916-024-03833-x

22. Matthews CE, Troiano RP, Salerno EA, et al. Exploration of Confounding Due to Poor Health in an Accelerometer-Mortality Study. Med Sci Sports Exerc. Dec 2020;52(12):2546–2553. doi:10.1249/mss.0000000000002405

23. Koemel NA, Ahmadi MN, Biswas RK, et al. Device-captured incidental physical activity, sedentary behaviour and all-cause mortality risk. Journal of Epidemiology and Community Health. 2025:jech-2024–222394. doi:10.1136/jech-2024-222394

24. Stamatakis E, Biswas RK, Koemel NA, et al. Dose Response of Incidental Physical Activity Against Cardiovascular Events and Mortality. Circulation. 2025/04/15 2025;151(15):1063–1075. doi:10.1161/CIRCULATIONAHA.124.072253

25. Pavey TG, Gilson ND, Gomersall SR, Clark B, Trost SG. Field evaluation of a random forest activity classifier for wrist-worn accelerometer data. Journal of Science and Medicine in Sport. 2017;20(1):75–80. doi:10.1016/j.jsams.2016.06.003

26. Ritz C, Baty F, Streibig JC, Gerhard D. Dose-Response Analysis Using R. PLOS ONE. 2016;10(12):e0146021. doi:10.1371/journal.pone.0146021

27. Ahmadi MN, Rezende LFM, Ferrari G, Del Pozo Cruz B, Lee IM, Stamatakis E. Do the associations of daily steps with mortality and incident cardiovascular disease differ by sedentary time levels? A device-based cohort study. British Journal of Sports Medicine. 2024;58(5):261. doi:10.1136/bjsports-2023-107221

28. Strain T, Wijndaele K, Dempsey PC, et al. Wearable-device-measured physical activity and future health risk. Nature Medicine. 2020/09/01 2020;26(9):1385–1391. doi:10.1038/s41591-020-1012-3

29. Mok A, Khaw K-T, Luben R, Wareham N, Brage S. Physical activity trajectories and mortality: population based cohort study. BMJ. 2019;365:l2323. doi:10.1136/bmj.l2323

30. Lu Y, Hajifathalian K, Ezzati M, Woodward M, Rimm EB, Danaei G. Metabolic mediators of the effects of body-mass index, overweight, and obesity on coronary heart disease and stroke: a pooled analysis of 97 prospective cohorts with 1.8 million participants. Lancet. 2014;383(9921):970–983. doi:10.1016/s0140-6736(13)61836-x

31. Krebs-Smith SM, Pannucci TE, Subar AF, et al. Update of the Healthy Eating Index: HEI-2015. J Acad Nutr Diet. Sep 2018;118(9):1591–1602. doi:10.1016/j.jand.2018.05.021

32. Hughes RA, Heron J, Sterne JAC, Tilling K. Accounting for missing data in statistical analyses: multiple imputation is not always the answer. International Journal of Epidemiology. 2019;48(4):1294–1304. doi:10.1093/ije/dyz032

33. Haneuse S, VanderWeele TJ, Arterburn D. Using the E-value to assess the potential effect of unmeasured confounding in observational studies. JAMA. 2019;321(6):602–603.

34. Saint-Maurice PF, Troiano RP, Bassett DR, et al. Association of daily step count and step intensity with mortality among US adults. Jama. 2020;323(12):1151–1160.

35. Inoue K, Tsugawa Y, Mayeda ER, Ritz B. Association of daily step patterns with mortality in US adults. JAMA Network Open. 2023;6(3):e235174–e235174.

36. Fishman EI, Steeves JA, Zipunnikov V, et al. Association between Objectively Measured Physical Activity and Mortality in NHANES. Med Sci Sports Exerc. Jul 2016;48(7):1303–11. doi:10.1249/mss.0000000000000885

37. Saint-Maurice PF, Graubard BI, Troiano RP, et al. Estimated Number of Deaths Prevented Through Increased Physical Activity Among US Adults. JAMA Internal Medicine. 2022;182(3):349–352. doi:10.1001/jamainternmed.2021.7755

38. Matthews CE, Keadle SK, Troiano RP, et al. Accelerometer-measured dose-response for physical activity, sedentary time, and mortality in US adults. The American journal of clinical nutrition. 2016;104(5):1424–1432.

39. Evenson KR, Wen F, Herring AH. Associations of accelerometry-assessed and self-reported physical activity and sedentary behavior with all-cause and cardiovascular mortality among US adults. American journal of epidemiology. 2016;184(9):621–632.

40. Schmid D, Ricci C, Leitzmann MF. Associations of objectively assessed physical activity and sedentary time with all-cause mortality in US adults: the NHANES study. PloS one. 2015;10(3):e0119591.

41. Saint-Maurice PF, Troiano RP, Matthews CE, Kraus WE. Moderate-to-Vigorous Physical Activity and All-Cause Mortality: Do Bouts Matter? Journal of the American Heart Association. 2018;7(6):e007678. doi:doi:10.1161/JAHA.117.007678

42. Ahmadi M, Koemel N, Biswas R, et al. Micropatterns of physical activity in relation to all-cause and cardiovascular disease mortality: the stealth lifestyle factor? medRxiv. 2024:2024.08.06.24311529. doi:10.1101/2024.08.06.24311529

43. Li Y, Xia P-F, Geng T-T, et al. Trends in Self-Reported Adherence to Healthy Lifestyle Behaviors Among US Adults, 1999 to March 2020. JAMA Network Open. 2023;6(7):e2323584–e2323584. doi:10.1001/jamanetworkopen.2023.23584

44. Batacan RB, Duncan MJ, Dalbo VJ, Tucker PS, Fenning AS. Effects of high-intensity interval training on cardiometabolic health: a systematic review and meta-analysis of intervention studies. British Journal of Sports Medicine. 2017;51(6):494. doi:10.1136/bjsports-2015-095841

45. Sultana RN, Sabag A, Keating SE, Johnson NA. The Effect of Low-Volume High-Intensity Interval Training on Body Composition and Cardiorespiratory Fitness: A Systematic Review and Meta-Analysis. Sports Medicine. 2019/11/01 2019;49(11):1687–1721. doi:10.1007/s40279-019-01167-w

46. Allison MK, Baglole JH, Martin BJ, Macinnis MJ, Gurd BJ, Gibala MJ. Brief Intense Stair Climbing Improves Cardiorespiratory Fitness. Medicine and science in sports and exercise. 2017;49(2):298–307. doi:10.1249/mss.0000000000001188

47. Adamson S, Kavaliauskas M, Yamagishi T, Phillips S, Lorimer R, Babraj J. Extremely short duration sprint interval training improves vascular health in older adults. Sport Sciences for Health. 2019/04/01 2019;15(1):123–131. doi:10.1007/s11332-018-0498-2

48. Ahmadi MN, Blodgett JM, Atkin AJ, et al. Relationship of device measured physical activity type and posture with cardiometabolic health markers: pooled dose– response associations from the Prospective Physical Activity, Sitting and Sleep Consortium. Diabetologia. 2024/06/01 2024;67(6):1051–1065. doi:10.1007/s00125-024-06090-y

49. Elgaddal N KE, Reuben C. Physical activity among adults aged 18 and over: United States, 2020. Journal Issue. 2022. https://stacks.cdc.gov/view/cdc/120213

50. van Hees VT, Sabia S, Jones SE, et al. Estimating sleep parameters using an accelerometer without sleep diary. Scientific reports. 2018;8(1):12975.

51. Ge C, Long B, Lu Q, Jiang Z, He Y. Associations of different type of physical activity with all-cause mortality in hypertension participants. Scientific Reports. 2024;14(1):7515. doi:10.1038/s41598-024-58197-2

52. Perez-Lasierra JL, Moreno-Franco B, González-Agüero A, Lobo E, Casajus JA. A cross-sectional analysis of the association between physical activity, depression, and all-cause mortality in Americans over 50 years old. Scientific Reports. 2022/02/10 2022;12(1):2264. doi:10.1038/s41598-022-05563-7

53. Porter AK, Cuthbertson CC, Evenson KR. Participation in specific leisure-time activities and mortality risk among U.S. adults. Annals of Epidemiology. 2020/10/01/ 2020;50:27–34.e1. 10.1016/j.annepidem.2020.06.006

54. Zhao G, Li C, Ford ES, et al. Leisure-time aerobic physical activity, muscle-strengthening activity and mortality risks among US adults: the NHANES linked mortality study. British Journal of Sports Medicine. 2014;48(3):244. doi:10.1136/bjsports-2013-092731

55. Tarp J, Hansen BH, Fagerland MW, Steene-Johannessen J, Anderssen SA, Ekelund U. Accelerometer-measured physical activity and sedentary time in a cohort of US adults followed for up to 13 years: the influence of removing early follow-up on associations with mortality. International Journal of Behavioral Nutrition and Physical Activity. 2020/03/14 2020;17(1):39. doi:10.1186/s12966-020-00945-4

