## Supplemental Figs 1-18, Supplemental Methods, Supplemental Table 1-7 for "Vigorous intermittent lifestyle physical activity (VILPA) and mortality risk among US adults: a wearables-based national cohort study"

### Table of Contents


| Page | Item |
|---|---|
| **4** | **Supplement Figure 1.** Flow diagram of participants in the study |
| **5** | **Supplement Figure 2.** VILPA frequency and hazard ratio with the relative rate of change for successive points |
| **6** | **Supplement Figure 3.** VILPA duration and hazard ratio with the relative rate of change for successive points |
| **7** | **Supplement Figure 4.** Survey adjusted absolute risk-based dose respon se curves of daily VILPA frequency (A) and duration (B) with all-cause mo rtality (n = 3,293; events = 290) |
| **8** | **Supplement Figure 5.** Survey adjusted dose response curves of daily VILPA frequency (A) and duration (B) with all-cause mortality (n = 2,390; events = 199) excluding those with frailty reported (current smoker, BMI < 18.5 and self-reported health condition as 'poor') |
| **9** | **Supplement Figure 6.** Survey adjusted dose response curves of daily VILPA frequency (A) and duration (B) with all-cause mortality (n = 2,731; events = 152) excluding previous CVD and cancer events |
| **10** | **Supplement Figure 7.** Survey adjusted dose response curves of daily VILPA frequency (A) and duration (B) with all-cause mortality (n = 2,987; events = 249) adjusted for cardiometabolic health markers |
| **11** | **Supplement Figure 8.** Survey adjusted dose response curves of daily VILPA frequency (A) and duration (B) with all-cause mortality (n = 3,293; events = 290) with additional adjustment for BMI |
| **12** | **Supplement Figure 9.** Survey adjusted dose response curves of daily VILPA with all-cause mortality (n = 3,070; events = 197) adjusted for the Healthy Eating Index 2015 |

| | |
|---|---|
| **13** | **Supplement Figure 10.** Survey adjusted dose response curves of daily VILPA bouts (A) and duration (B) with all-cause mortality (n = 3,164; events = 231) excluding samples with VILPA = 0 min/day |
| **14** | **Supplement Figure 11.** Survey adjusted dose response curves of daily VILPA frequency (A) and duration (B) with all-cause mortality (n = 2,202; events = 87) excluding samples with VILPA < 0.5 min/day |
| **15** | **Supplement Figure 12.** Survey adjusted dose response curves of daily VILPA frequency (A) and duration (B) with all-cause mortality (n = 3,210; events = 289) with VILPA maximum truncated at 97.5th percentile (8.9 mins/day) |
| **16** | **Supplement Figure 13.** Survey adjusted dose response curves of daily VILPA frequency (A) and duration (B) with all-cause mortality after considering deaths from accidents/unintentional injuries as competing interests (n = 3,293; events = 279, competing events = 11) |
| **17** | **Supplement Figure 14.** Survey adjusted dose response curves of daily VILPA frequency (A) and duration (B) with all-cause mortality after considering deaths from residual causes as competing interests (n = 3,293; events = 222, competing events = 68) |
| **18** | **Supplement Figure 15.** Survey adjusted dose response curves of daily VILPA frequency (A) and duration (B) with all-cause mortality (n = 3,293; events = 290) using the median as the referent point |
| **19** | **Supplement Figure 16.** Survey adjusted dose response curves of daily VILPA with all-cause mortality (n = 5,293; events = 417) alternative non-exerciser definition |
| **20** | **Supplement Figure 17.** Survey adjusted dose response curves of daily VILPA with all-cause mortality (n = 3,738; events = 301) alternative non-exerciser definition |
| **21** | **Supplement Figure 18.** Survey adjusted dose response curves of daily VILPA frequency (A) and duration (B) with all-cause mortality after removing samples with missing data (n = 2,592; events = 235) |
| **22** | **Supplement Methods.** Additional study design details and wearable behaviour classification methods |

| | |
|---|---|
| **25** | **Supplement Table 1.** Questions to determine leisure time physical activity |
| **26** | **Supplement Table 2.** Mortality definitions and ICD-10 Codes |
| **28** | **Supplement Table 3.** Covariate definitions |
| **30** | **Supplement Table 4.** Model variance inflation factors |
| **32** | **Supplement Table 5.** Interaction test between sex and VILPA with all-cause mortality |
| **33** | **Supplement Table 6.** E-values for VILPA frequency and duration |
| **34** | **Supplement Table 7.** STROBE statement |

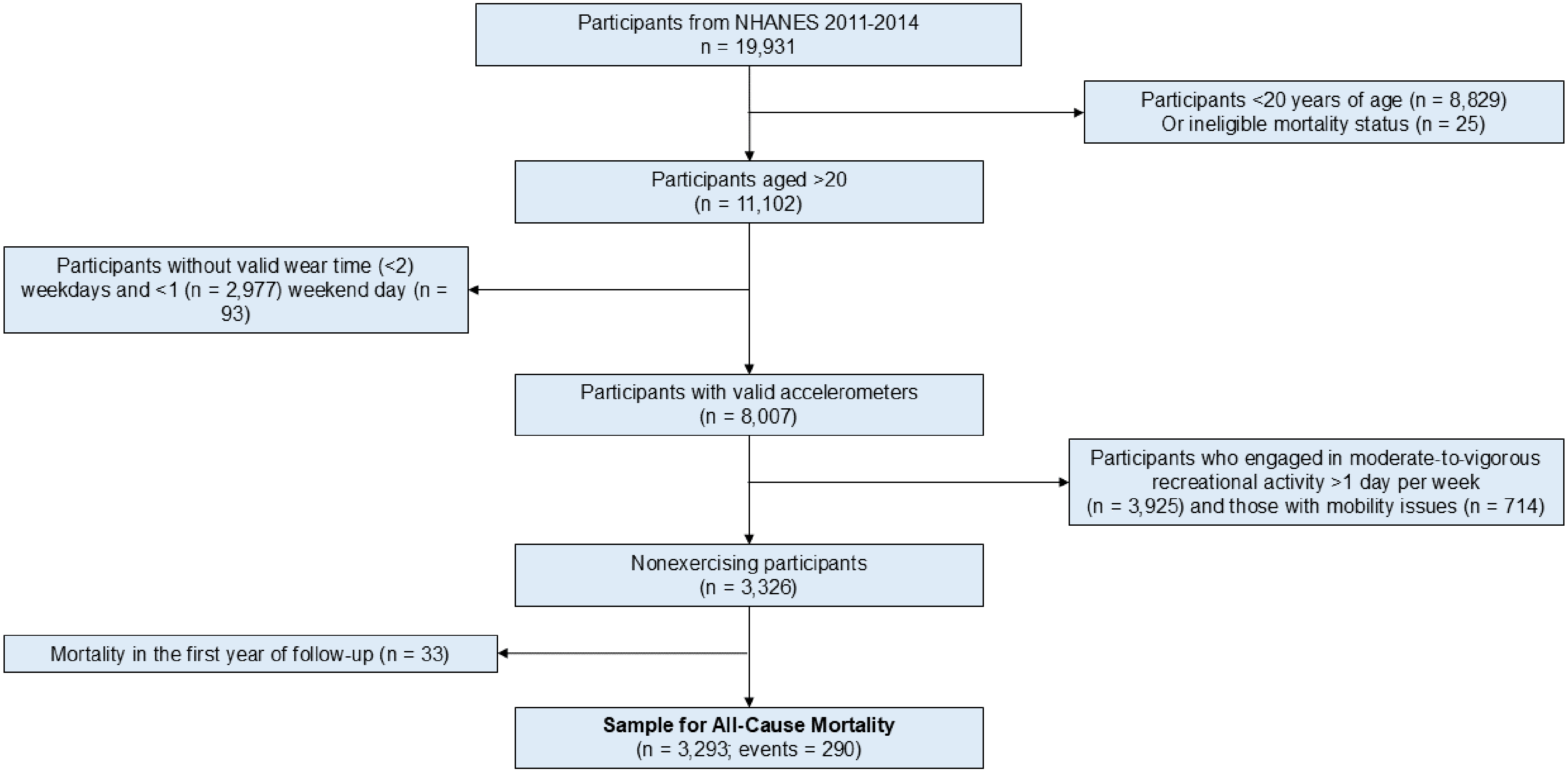


**Supplement Figure 1.** Flow diagram of participants in the study

**Supplement Figure 2.** VILPA frequency and hazard ratio with the relative rate of change for successive points.

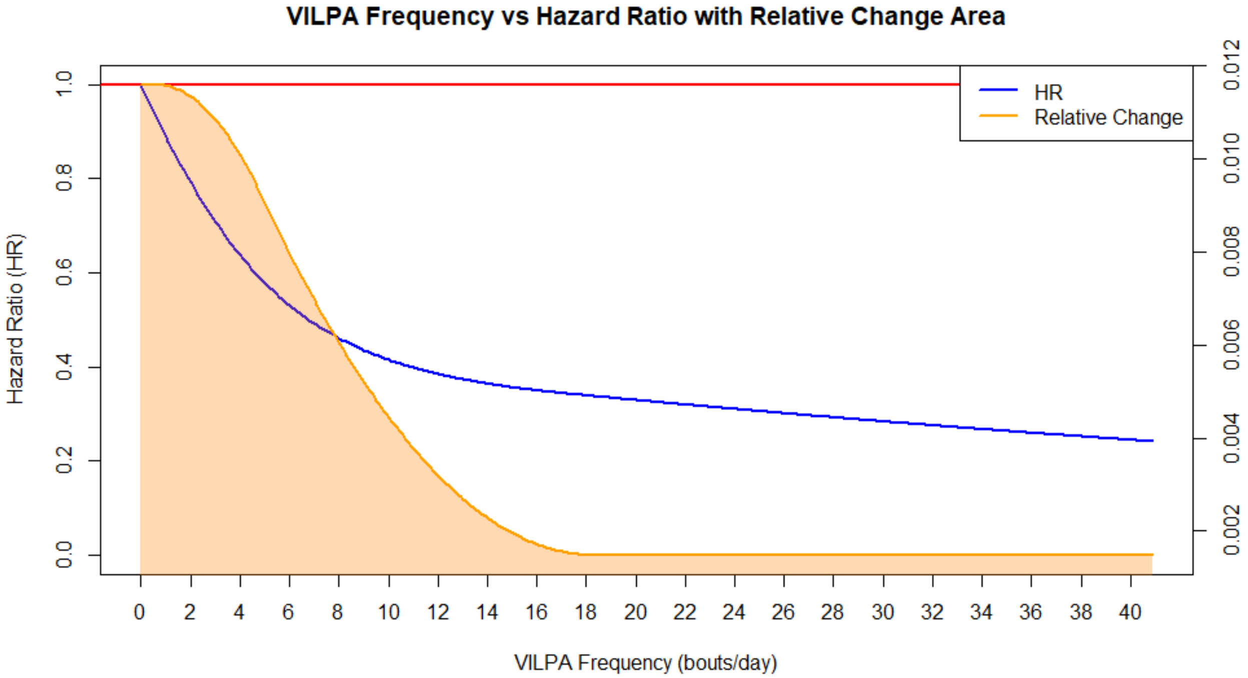


**Legend**: The plot shows the dose-response hazard ratio for all-cause mortality (blue line) and the relative change between successive points as the orange line. The intersection of these two points is considered the point of diminishing returns. Analyses were adjusted for sex, age, income, education, ethnicity, fruit and vegetable consumption, smoking history, total physical activity energy expenditure, total daily VILPA duration, alcohol consumption, sleep duration, duration of light and moderate intensity, discretionary screentime, medication use (glycaemic control, blood pressure, cholesterol), family history of diabetes and CVD, and previous history of CVD and diabetes.

**Supplement Figure 3.** VILPA duration and hazard ratio with the relative rate of change for successive points.

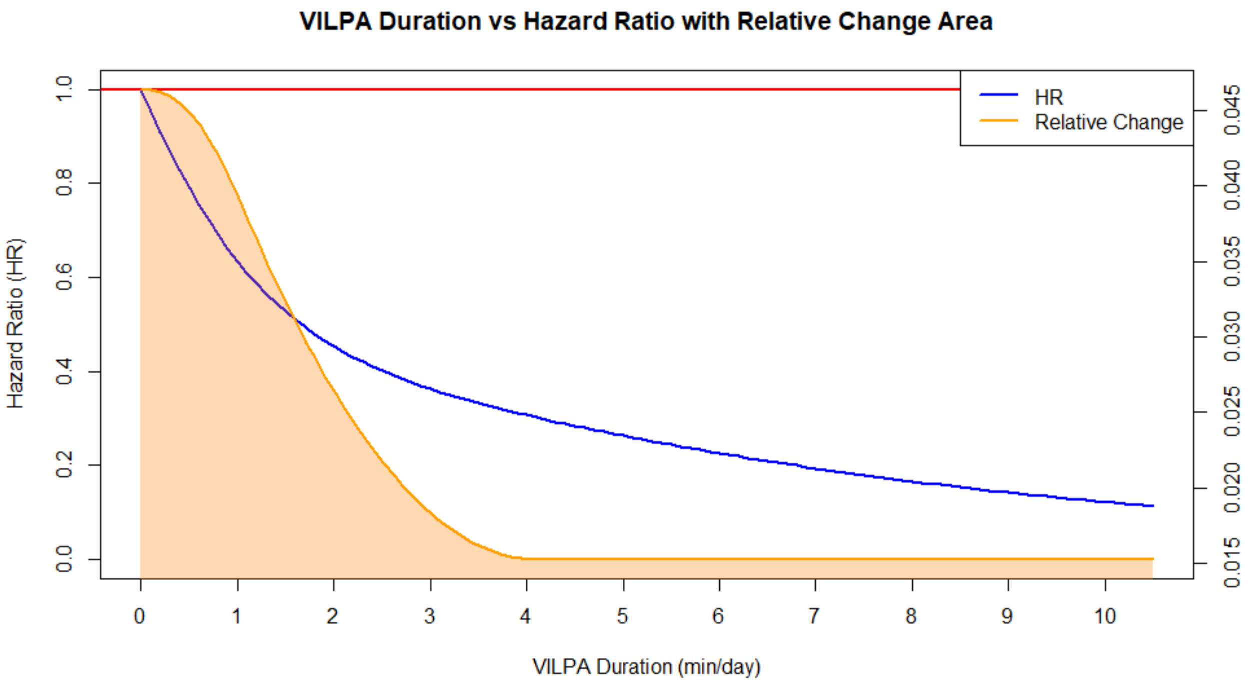


**Legend**: The plot shows the dose-response hazard ratio for all-cause mortality (blue line) and the relative change between successive points as the orange line. The intersection of these two points is considered the point of diminishing returns. Analyses were adjusted for sex, age, income, education, ethnicity, fruit and vegetable consumption, smoking history, physical activity energy expenditure from LPA and MPA, VILPA bouts over 1-minute, alcohol consumption, sleep duration, duration of light and moderate intensity, discretionary screentime, medication use (glycaemic control, blood pressure, cholesterol), family history of diabetes and CVD, and previous history of CVD and diabetes.

**Supplement Figure 4.** Survey adjusted absolute risk-based dose response curves o f daily VILPA frequency (A) and duration (B) with all-cause mortality (n = 3,293; event s = 290).

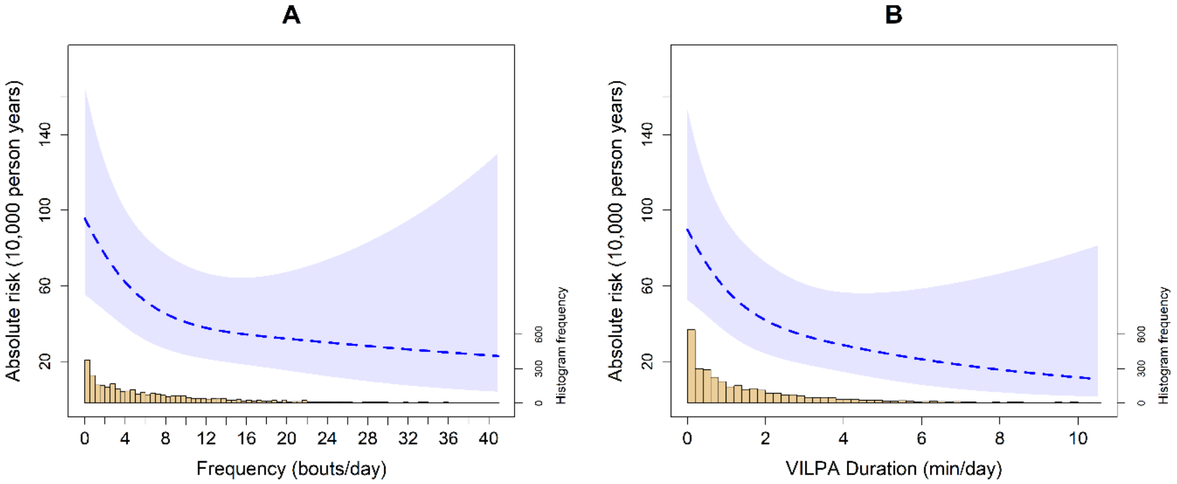


**Legend***:* Analyses were adjusted for sex, age, income, education, ethnicity, fruit and vegetable consumption, smoking history, physical activity energy expenditure from L PA and MPA, VILPA bouts over 1-minute, alcohol consumption, sleep duration, durati on of light and moderate intensity, discretionary screentime, medication use (glycaem ic control, blood pressure, cholesterol), family history of diabetes and CVD, and previ ous history of CVD, diabetes, and cancer. VILPA bout (A) was further adjusted by ene rgy expenditure by vigorous intensity and VILPA duration (B) was further adjusted for VILPA bouts over 1-minute bout. All analyses excluded participants who had an even t in the first year of follow-up. All analyses were adjusted for strata, cluster, and surve y weights.

**Supplement Figure 5:** Survey adjusted dose response curves of daily VILPA bouts (A) and duration (B) with all-cause mortality (n = 2,390; events = 199) excluding those with frailty reported (current smoker, BMI < 18.5 and self-reported health condition as ‘poor’).

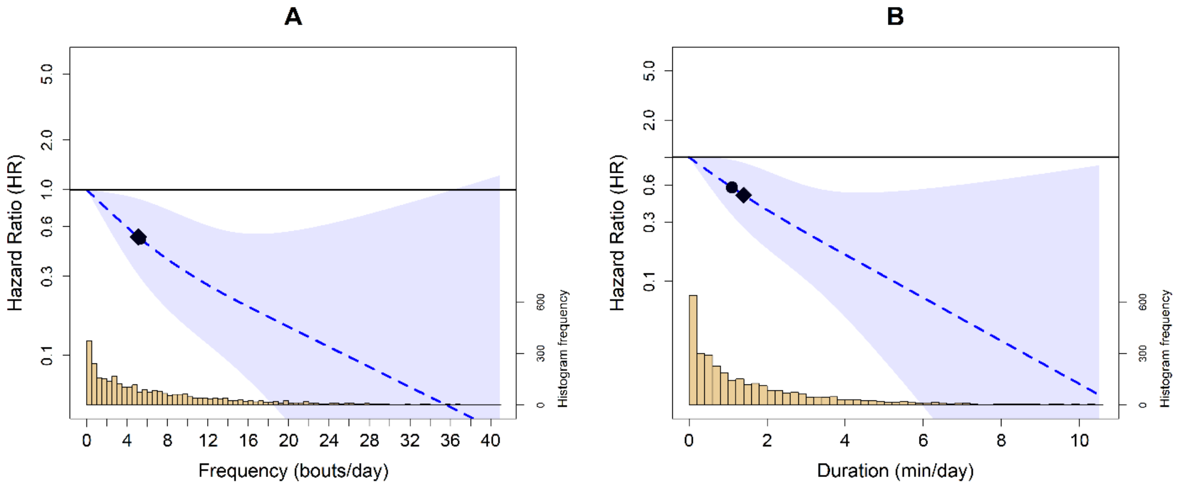


**Legend:** Analyses were adjusted for sex, age, income, education, ethnicity, fruit and vegetable consumption, smoking history, physical activity energy expenditure from LPA and MPA, alcohol consumption, sleep duration, energy expenditure by light and moderate intensity, discretionary screentime, medication use (glycaemic control, blood pressure, cholesterol), family history of diabetes and CVD, and previous history of CVD, diabetes, and cancer. VILPA bout (A) was further adjusted by energy expenditure by vigorous intensity and VILPA duration (B) was further adjusted for VILPA bouts over 1-minute bout. All analyses excluded participants who had an event in the first year of follow-up. All analyses were adjusted for strata, cluster, and survey weights. Reference was set to zero for VILPA frequency and duration. Diamond, minimal dose, as indicated by the ED50 statistic which estimates the daily duration of VILPA associated with 50% of optimal risk reduction. Circle, HR associated with the median VILPA value.

**Supplement Figure 6:** Survey adjusted dose response curves of daily VILPA bouts (A) and duration (B) with all-cause mortality (n = 2,731; events = 152) excluding previous CVD and cancer events.

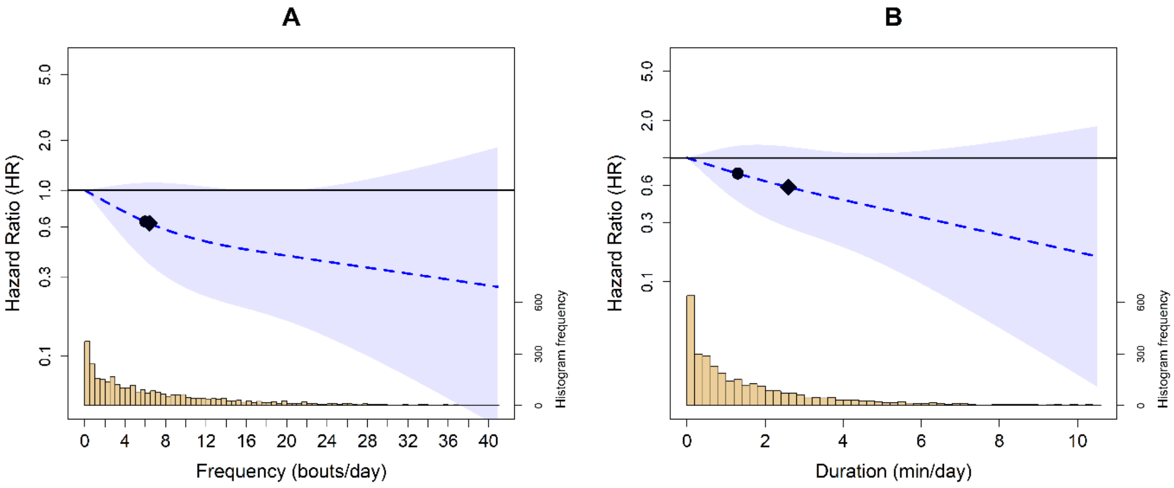


**Legend:** Analyses were adjusted for sex, age, income, education, ethnicity, fruit and vegetable consumption, smoking history, physical activity energy expenditure from LPA and MPA, alcohol consumption, sleep duration, energy expenditure by light and moderate intensity, discretionary screentime, medication use (glycaemic control, blood pressure, cholesterol), family history of diabetes and CVD, and previous history of CVD, diabetes, and cancer. VILPA bout (A) was further adjusted by energy expenditure by vigorous intensity and VILPA duration (B) was further adjusted for VILPA bouts over 1-minute bout. All analyses were adjusted for strata, cluster, and survey weights. Reference was set to zero for VILPA frequency and duration. Diamond, minimal dose, as indicated by the ED50 statistic which estimates the daily duration of VILPA associated with 50% of optimal risk reduction. Circle, HR associated with the median VILPA value.

**Supplement Figure 7:** Survey adjusted dose response curves of daily VILPA bouts (A) and duration (B) with all-cause mortality (n = 2,987; events = 249) adjusted for cardiometabolic health markers.

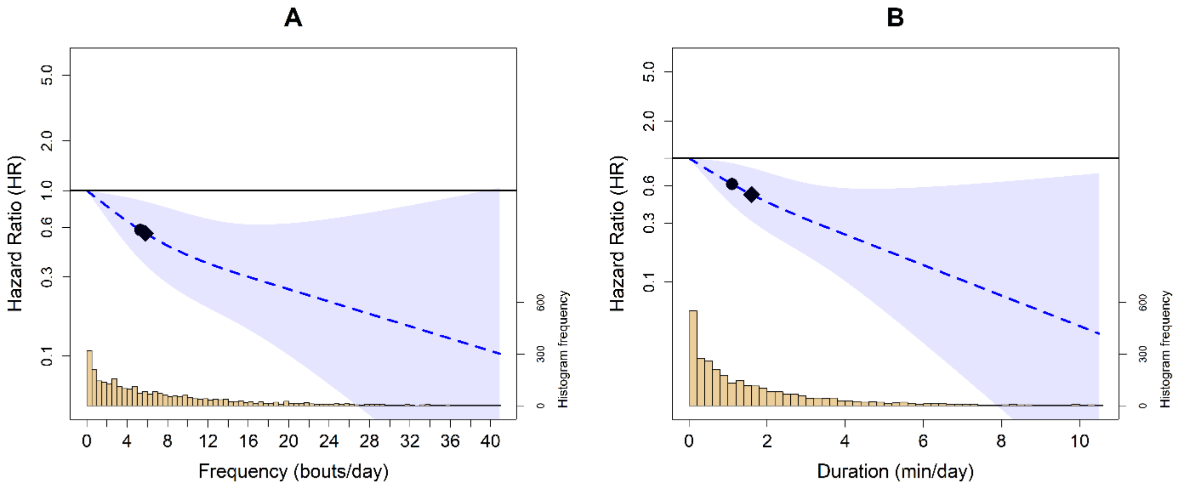


**Legend:** Analyses were adjusted for sex, age, income, education, ethnicity, fruit and vegetable consumption, smoking history, physical activity energy expenditure from LPA and MPA, alcohol consumption, waist circumference, fasting blood glucose, HDL cholesterol, total cholesterol, systolic blood pressure, diastolic blood pressure, sleep duration, energy expenditure by light and moderate intensity, discretionary screentime, medication use (glycaemic control, blood pressure, cholesterol), family history of diabetes and CVD, and previous history of CVD, diabetes, and cancer. VILPA bout (A) was further adjusted by energy expenditure by vigorous intensity and VILPA duration (B) was further adjusted for VILPA bouts over 1-minute bout. All analyses were adjusted for strata, cluster, and survey weights. Reference was set to zero for VILPA frequency and duration. Diamond, minimal dose, as indicated by the ED50 statistic which estimates the daily duration of VILPA associated with 50% of optimal risk reduction. Circle, HR associated with the median VILPA value.

**Supplement Figure 8:** Survey adjusted dose response curves of daily VILPA bouts (A) and duration (B) with all-cause mortality (n = 3,293; events = 290) with additional adjustment for BMI.

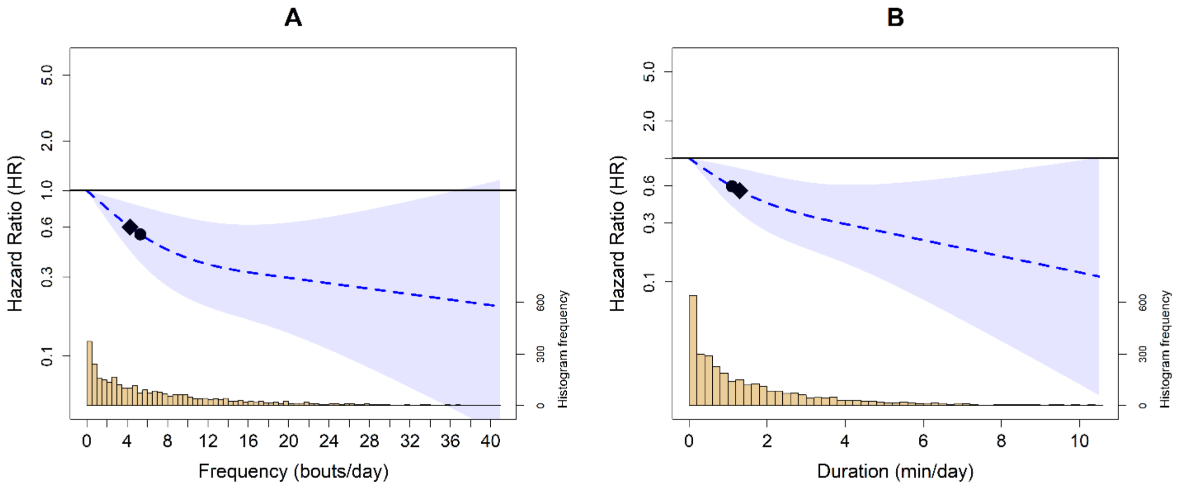


**Legend:** Analyses were adjusted for sex, age, income, education, ethnicity, fruit and vegetable consumption, smoking history, physical activity energy expenditure from LPA and MPA, alcohol consumption, sleep duration, energy expenditure by light and moderate intensity, discretionary screentime, medication use (glycaemic control, blood pressure, cholesterol), family history of diabetes and CVD, BMI, and previous history of CVD, diabetes, and cancer. VILPA bout (A) was further adjusted by energy expenditure by vigorous intensity and VILPA duration (B) was further adjusted for VILPA bouts over 1-minute bout. All analyses excluded participants who had an event in the first year of follow-up. All analyses were adjusted for strata, cluster, and survey weights. Reference was set to zero for VILPA frequency and duration. Diamond, minimal dose, as indicated by the ED50 statistic which estimates the daily duration of VILPA associated with 50% of optimal risk reduction. Circle, HR associated with the median VILPA value.

**Supplement Figure 9:** Survey adjusted dose response curves of daily VILPA with all-cause mortality (n = 3,070; events = 197) adjusted for the Healthy Eating Index 2015.

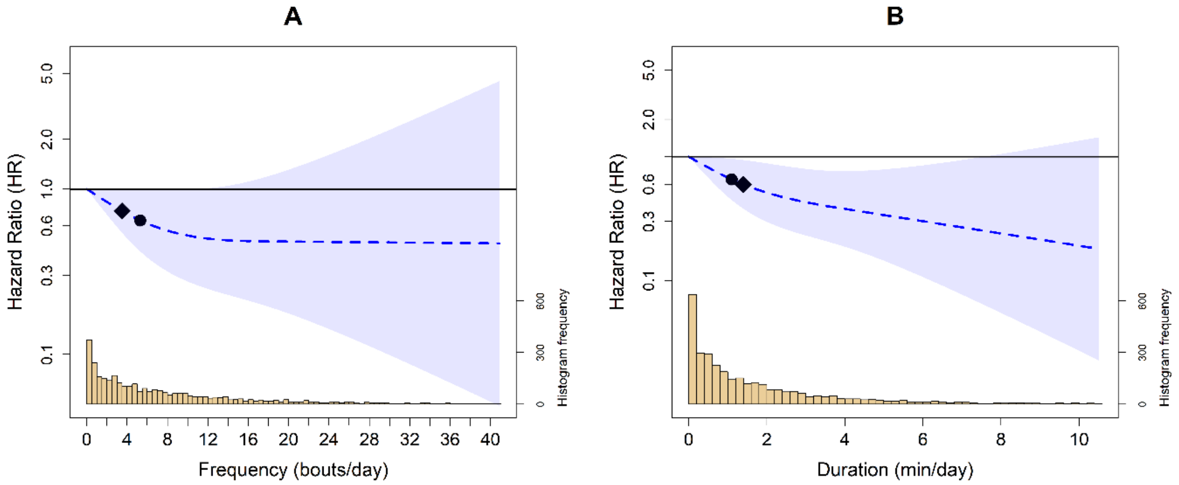


**Legend:** Analyses were adjusted for sex, age, income, education, ethnicity, Healthy Eating Index 2015, smoking history, physical activity energy expenditure from LPA and MPA, VILPA bouts over 1-minute, alcohol consumption, sleep duration, duration of light and moderate intensity, discretionary screentime, medication use (glycaemic control, blood pressure, cholesterol), family history of diabetes and CVD, and previous history of CVD, diabetes, and cancer, waist circumference, total cholesterol, HbA1c, diastolic and systolic blood pressure. All analyses excluded participants who had an event in the first year of follow-up. All analyses were adjusted for strata, cluster, and survey weights. Reference was set to zero for VILPA frequency and duration. Diamond, minimal dose, as indicated by the ED50 statistic which estimates the daily duration of VILPA associated with 50% of optimal risk reduction. Circle, HR is associated with the median VILPA value.

**Supplement Figure 10:** Survey adjusted dose response curves of daily VILPA bouts (A) and duration (B) with all-cause mortality (n = 3,164; events = 231) excluding samples with VILPA = 0 min/day.

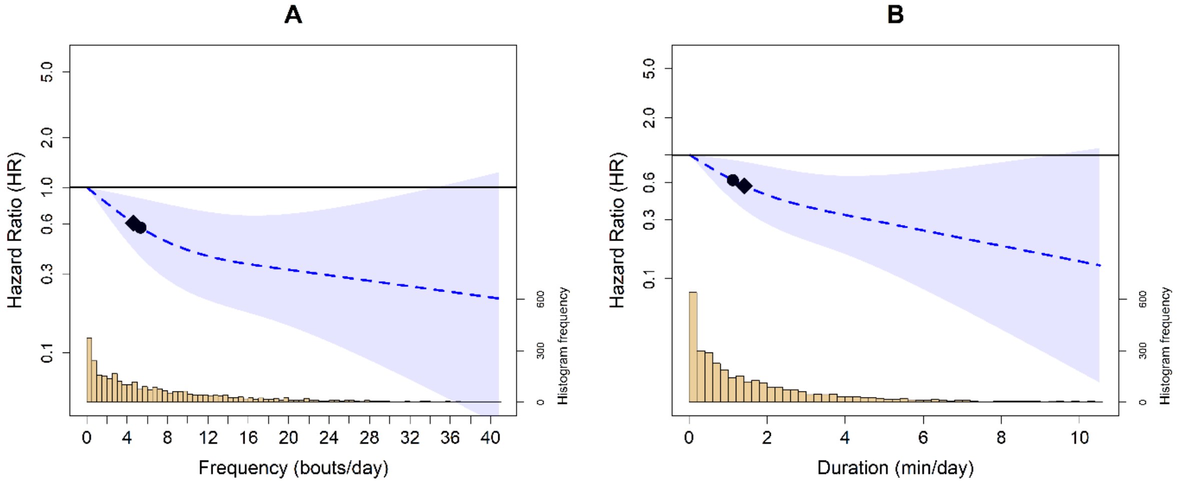


**Legend:** Analyses were adjusted for sex, age, income, education, ethnicity, fruit and vegetable consumption, smoking history, physical activity energy expenditure from LPA and MPA, alcohol consumption, sleep duration, energy expenditure by light and moderate intensity, discretionary screentime, medication use (glycaemic control, blood pressure, cholesterol), family history of diabetes and CVD, and previous history of CVD, diabetes, and cancer. VILPA bout (A) was further adjusted by energy expenditure by vigorous intensity and VILPA duration (B) was further adjusted for VILPA bouts over 1-minute bout. All analyses excluded participants who had an event in the first year of follow-up. All analyses were adjusted for strata, cluster, and survey weights. Reference was set to 0.1 for VILPA duration. Diamond, minimal dose, as indicated by the ED50 statistic which estimates the daily duration of VILPA associated with 50% of optimal risk reduction. Circle, HR associated with the median VILPA value.

**Supplement Figure 11:** Survey adjusted dose response curves of daily VILPA bouts (A) and duration (B) with all-cause mortality (n = 2,202; events = 87) excluding samples with VILPA < 0.5 min/day.

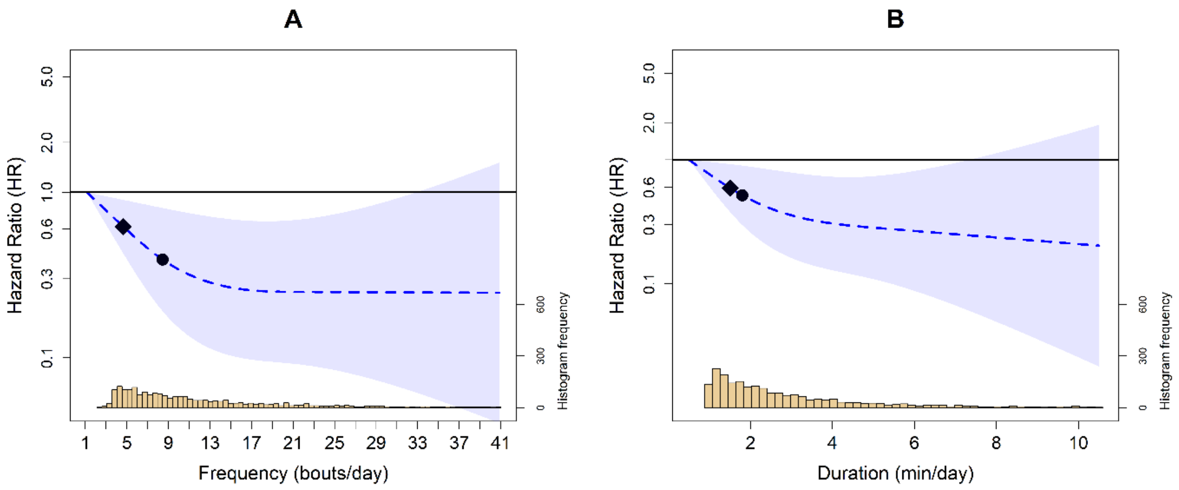


**Legend:** Analyses were adjusted for sex, age, income, education, ethnicity, fruit and vegetable consumption, smoking history, physical activity energy expenditure from LPA and MPA, alcohol consumption, sleep duration, energy expenditure by light and moderate intensity, discretionary screentime, medication use (glycaemic control, blood pressure, cholesterol), family history of diabetes and CVD, and previous history of CVD, diabetes, and cancer. VILPA bout (A) was further adjusted by energy expenditure by vigorous intensity and VILPA duration (B) was further adjusted for VILPA bouts over 1-minute bout. All analyses were adjusted for strata, cluster, and survey weights. Reference was set to 0.5 for VILPA duration. Diamond, minimal dose, as indicated by the ED50 statistic which estimates the daily duration of VILPA associated with 50% of optimal risk reduction. Circle, HR associated with the median VILPA value.

**Supplement Figure 12:** Survey adjusted dose response curves of daily VILPA bouts (A) and duration (B) with all-cause mortality (n = 3,210; events = 289) with VILPA maximum truncated at 97.5th percentile (8.9 mins/day).

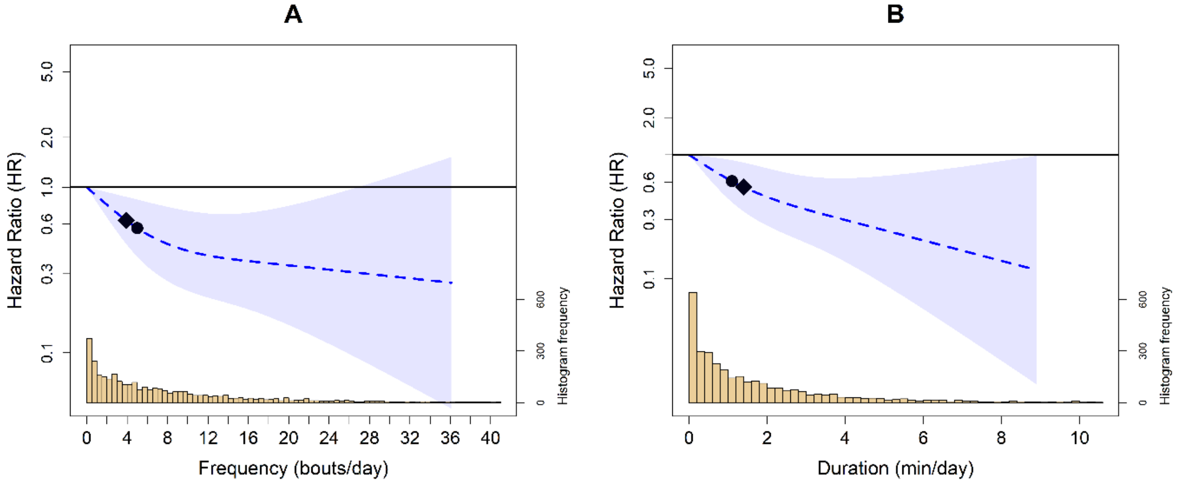


**Legend:** Analyses were adjusted for sex, age, income, education, ethnicity, fruit and vegetable consumption, smoking history, physical activity energy expenditure from LPA and MPA, alcohol consumption, sleep duration, energy expenditure by light and moderate intensity, discretionary screentime, medication use (glycaemic control, blood pressure, cholesterol), family history of diabetes and CVD, and previous history of CVD, diabetes, and cancer. VILPA bout (A) was further adjusted by energy expenditure by vigorous intensity and VILPA duration (B) was further adjusted for VILPA bouts over 1-minute bout. All analyses were adjusted for strata, cluster, and survey weights. Reference was set to zero for VILPA frequency and duration. Diamond, minimal dose, as indicated by the ED50 statistic which estimates the daily duration of VILPA associated with 50% of optimal risk reduction. Circle, HR associated with the median VILPA value.

**Supplement Figure 13:** Survey adjusted dose response curves of daily VILPA bouts (A) and duration (B) with all-cause mortality after considering deaths from accidents/unintentional injuries as competing interests (n = 3,293; events = 279, competing events = 11).

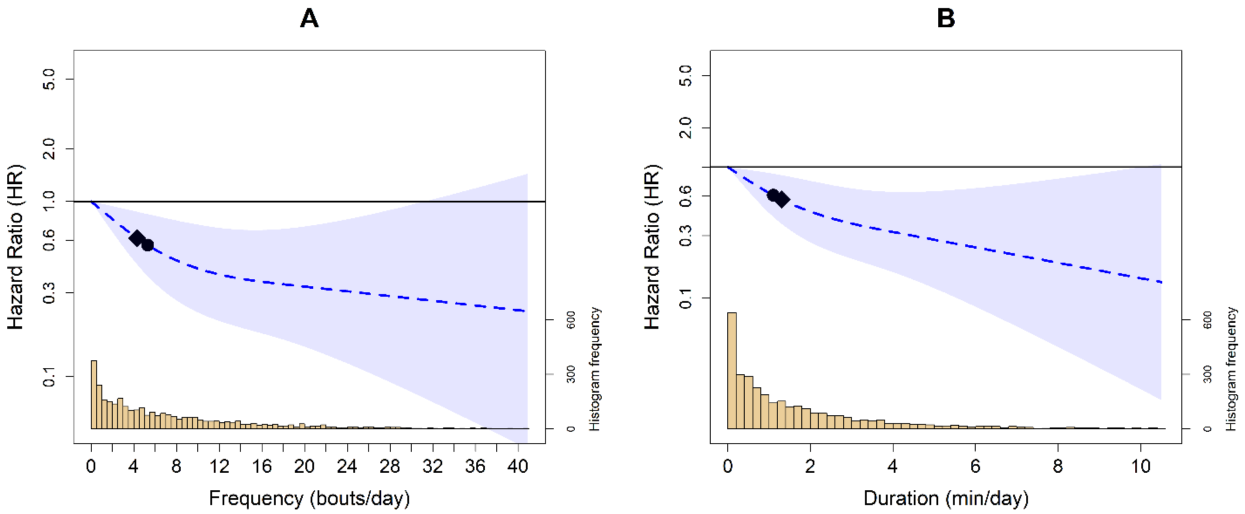


**Legend:** Analyses were adjusted for sex, age, income, education, ethnicity, fruit and vegetable consumption, smoking history, physical activity energy expenditure from LPA and MPA, alcohol consumption, sleep duration, energy expenditure by light and moderate intensity, discretionary screentime, medication use (glycaemic control, blood pressure, cholesterol), family history of diabetes and CVD, and previous history of CVD, diabetes, and cancer. VILPA bout (A) was further adjusted by energy expenditure by vigorous intensity and VILPA duration (B) was further adjusted for VILPA bouts over 1-minute bout. All analyses excluded participants who had an event in the first year of follow-up. All analyses were adjusted for strata, cluster, and survey weights. Reference was set to zero for VILPA frequency and duration. Diamond, minimal dose, as indicated by the ED50 statistic which estimates the daily duration of VILPA associated with 50% of optimal risk reduction. Circle, HR associated with the median VILPA value.

**Supplement Figure 14:** Survey adjusted dose response curves of daily VILPA bouts (A) and duration (B) with all-cause mortality after considering deaths from residual causes as competing interests (n = 3,293; events = 222, competing events = 68).

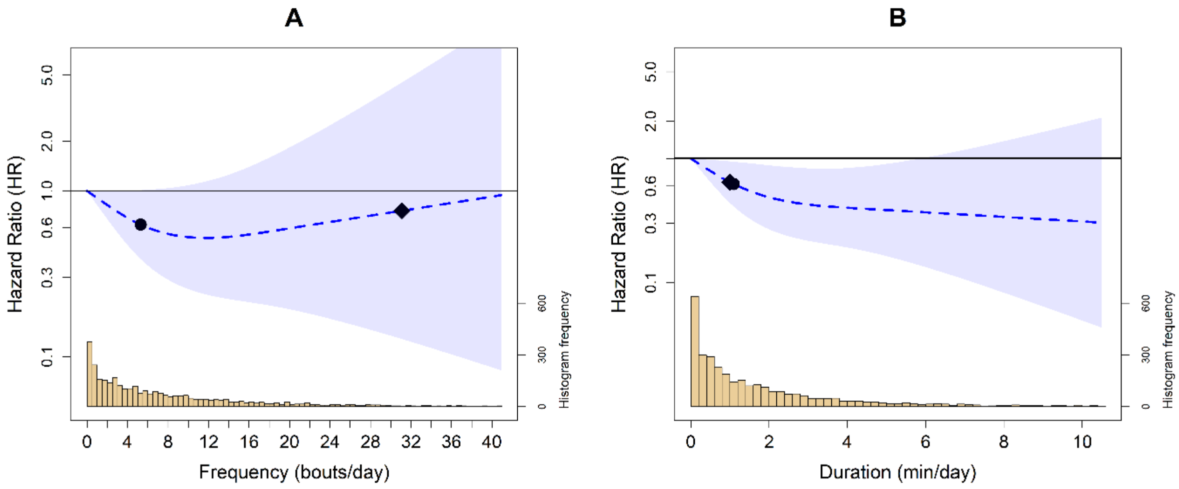


**Legend:** Analyses were adjusted for sex, age, income, education, ethnicity, fruit and vegetable consumption, smoking history, physical activity energy expenditure from LPA and MPA, alcohol consumption, sleep duration, energy expenditure by light and moderate intensity, discretionary screentime, medication use (glycaemic control, blood pressure, cholesterol), family history of diabetes and CVD, and previous history of CVD, diabetes, and cancer. VILPA bout (A) was further adjusted by energy expenditure by vigorous intensity and VILPA duration (B) was further adjusted for VILPA bouts over 1-minute bout. All analyses excluded participants who had an event in the first year of follow-up. All analyses were adjusted for strata, cluster, and survey weights. Reference was set to zero for VILPA frequency and duration. Diamond, minimal dose, as indicated by the ED50 statistic which estimates the daily duration of VILPA associated with 50% of optimal risk reduction. Circle, HR associated with the median VILPA value.

**Supplement Figure 15:** Survey adjusted dose response curves of daily VILPA bouts (A) and duration (B) with all-cause mortality (n = 3,293; events = 290) using the median as the referent point

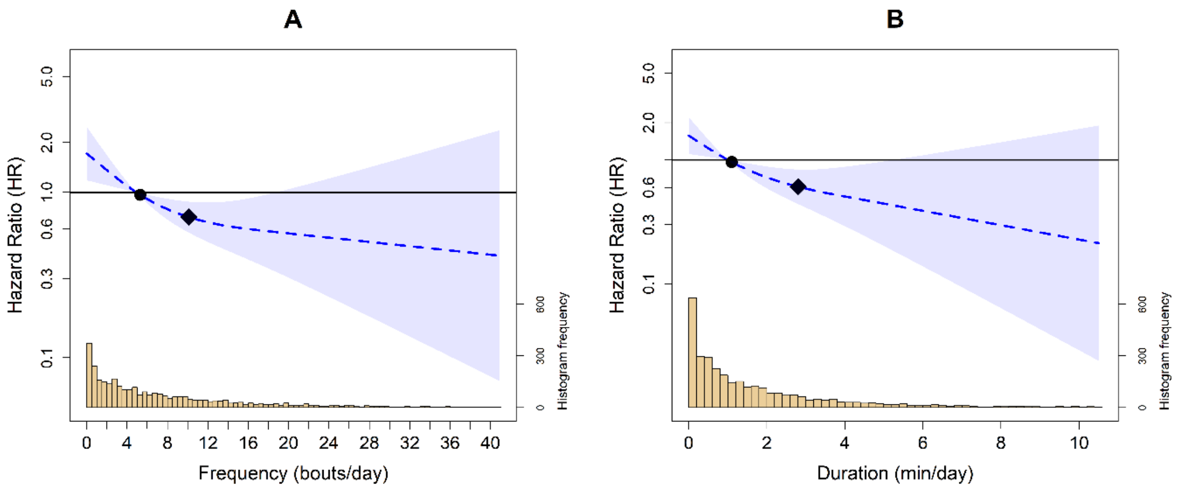


**Legend:** Analyses were adjusted for sex, age, income, education, ethnicity, fruit and vegetable consumption, smoking history, physical activity energy expenditure from LPA and MPA, alcohol consumption, sleep duration, energy expenditure by light and moderate intensity, discretionary screentime, medication use (glycaemic control, blood pressure, cholesterol), family history of diabetes and CVD, and previous history of CVD, diabetes, and cancer. VILPA bout (A) was further adjusted by energy expenditure by vigorous intensity and VILPA duration (B) was further adjusted for VILPA bouts over 1-minute bout. All analyses excluded participants who had an event in the first year of follow-up. All analyses were adjusted for strata, cluster, and survey weights. Reference was set to 5.3 for VILPA bouts per day and 1.1 minutes per day for VILPA duration. Diamond, minimal dose, as indicated by the ED50 statistic which estimates the daily duration of VILPA associated with 50% of optimal risk reduction. Circle, HR associated with the median VILPA value.

**Supplement Figure 16:** Survey adjusted dose response curves of daily VILPA with all-cause mortality (n = 5,293; events = 417) alternative non-exerciser definition.

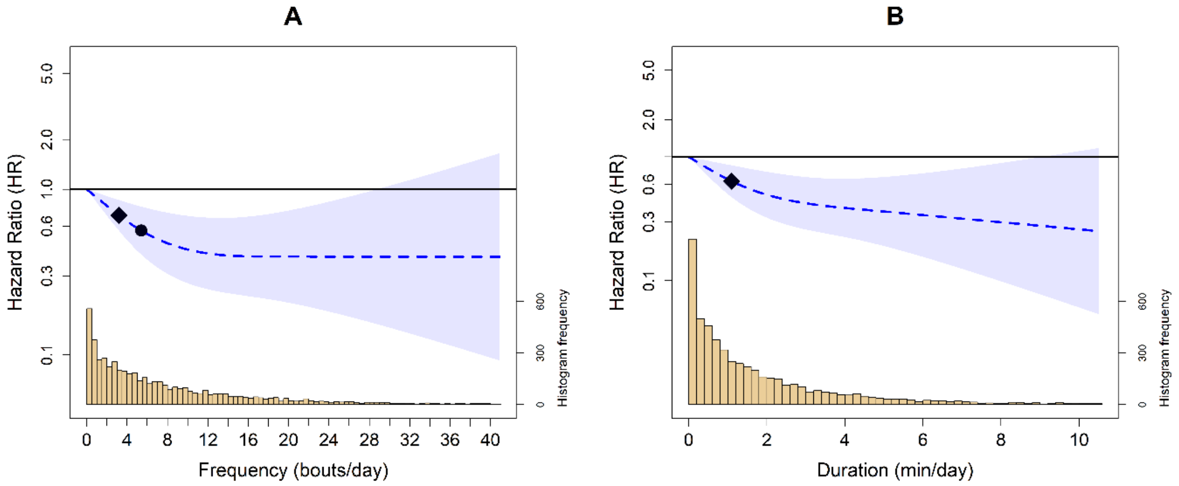


**Legend:** Model included non-exerciser using an alternative GPAQ definition including adults who reported only vigorous recreational exercise. Analyses were adjusted for sex, age, income, education, ethnicity, fruit and vegetable consumption, smoking history, physical activity energy expenditure from LPA and MPA, VILPA bouts over 1-minute, alcohol consumption, sleep duration, duration of light and moderate intensity, discretionary screentime, medication use (glycaemic control, blood pressure, cholesterol), family history of diabetes and CVD, and previous history of CVD, diabetes, and cancer. All analyses excluded participants who had an event in the first year of follow-up. All analyses were adjusted for strata, cluster, and survey weights. Reference was set to 0 for VILPA frequency and duration. Diamond, minimal dose, as indicated by the ED50 statistic which estimates the daily duration of VILPA associated with 50% of optimal risk reduction. Circle, HR is associated with the median VILPA value.

**Supplement Figure 17:** Survey adjusted dose response curves of daily VILPA with all-cause mortality (n = 3,738; events = 301) alternative non-exerciser definition.

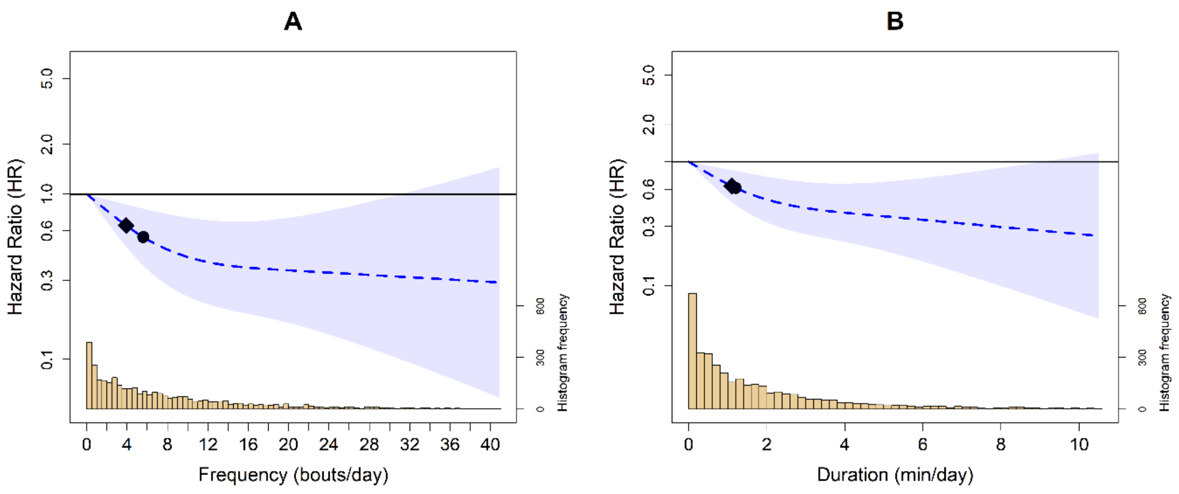


**Legend:** Model included non-exerciser using an alternative GPAQ definition including adults who reported ≤1 day per week of recreational moderate to vigorous physical activity per week. Analyses were adjusted for sex, age, income, education, ethnicity, fruit and vegetable consumption, smoking history, physical activity energy expenditure from LPA and MPA, VILPA bouts over 1-minute, alcohol consumption, sleep duration, duration of light and moderate intensity, discretionary screentime, medication use (glycaemic control, blood pressure, cholesterol), family history of diabetes and CVD, and previous history of CVD, diabetes, and cancer. All analyses excluded participants who had an event in the first year of follow-up. All analyses were adjusted for strata, cluster, and survey weights. Reference was set to 0 for VILPA frequency and daily duration. Diamond, minimal dose, as indicated by the ED50 statistic which estimates the daily duration of VILPA associated with 50% of optimal risk reduction. Circle, HR is associated with the median VILPA value.

**Supplement Figure 18:** Survey adjusted dose response curves of daily VILPA frequency (A) and duration (B) with all-cause mortality after removing samples with missing data (n = 2,592; events = 235).

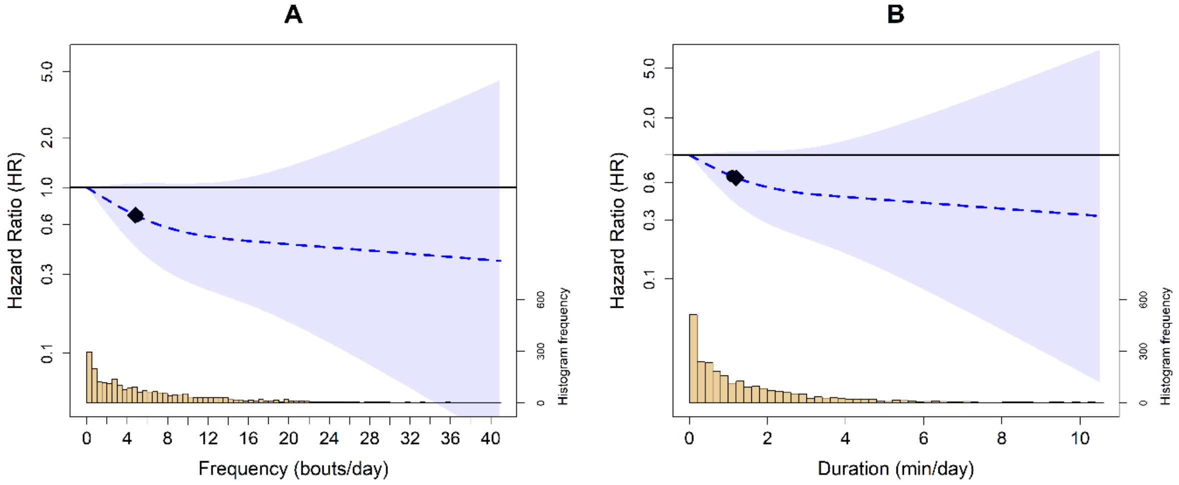


**Legend:** Analyses were adjusted for sex, age, income, education, ethnicity, fruit and vegetable consumption, smoking history, physical activity energy expenditure from LPA and MPA, alcohol consumption, sleep duration, energy expenditure by light and moderate intensity, discretionary screentime, medication use (glycaemic control, blood pressure, cholesterol), family history of diabetes and CVD, and previous history of CVD, diabetes, and cancer. VILPA frequency (A) was further adjusted by energy expenditure by vigorous intensity and VILPA duration (B) was further adjusted for VILPA bouts over 1-minute bout. All analyses excluded participants who had an event in the first year of follow-up. All analyses were adjusted for strata, cluster, and survey weights. Reference was set to zero for VILPA frequency and duration. Diamond, minimal dose, as indicated by the ED50 statistic which estimates the daily duration of VILPA associated with 50% of optimal risk reduction. Circle, HR associated with the median VILPA value.

**Supplement Methods**. Additional study design details and wearable behaviour classification methods

### VILPA Definition and Bout Length Selection

The definition of VILPA and selection of bout length is based on previously collected laboratory data[1]. In this study, participants completed a range of five activities while wearing an indirect calorimetry unit (Cosmed K5) and Polar heart rate monitor. These activities included (1) walking on a flat surface at a self-selected ‘very fast’ pace; (2) walking on a flat surface while carrying shopping-like bags equivalent to 5% of body weight at a self-defined ‘fast’ pace; (3) walking on a flat surface while carrying shopping-like bags equivalent to 10% of body weight at a self-defined ‘fast’ pace; (4) walking at a 2.5% gradient at a self-defined ‘very fast’ pace (treadmill); and (5) walking at a 7.0% gradient at a self-defined ‘very fast’ pace (treadmill). The participants completed these activities until reaching vigorous intensity measured by reaching two of three criteria: (1) %VO2max (percentage of maximal oxygen uptake) (≥64%); (2) %HRmax (percentage of maximal heart rate) (≥77%); and (3) rating of perceived exertion (Borg scale) ≥15. For %VO2max and %HRmax, the threshold had to be met for at least 30 consecutive seconds to minimize the effects of noise. Between activities, participants had 5 min of seated recovery, or until heart rate and breathing returned to resting levels. Resting VO2 and heart rate were measured at the beginning of each session with the participant lying supine using 5 min of steady-state (coefficient of variation ≤ 10%). As the mean time required to reach vigorous intensity in two of the above three physiological intensity indices was 73.5 s (s.d. 26.2 s) across all activities we chose to use 1-minute VILPA our bout length duration[2,3]. The frequency of raw bouts per day was defined as the number of occasions where the vigorous activity was completed in a bout of 10s up to 1 minute. Bouts occur as whole integers, however, for this study we report the sample median of bouts.

### Wearable physical activity intensity and posture classification

Incidental physical activity was classified using a validated two-stage random forest activity classifier that first classifies each 10 second window (epoch) as sedentary (lying or sitting still), standing utilitarian (for example, ironing a shirt, washing dishes)), walking (for example, gardening, active commuting, mopping floors), or running/high energetic activities (for example, active playing with children) (**Diagram A**)[4-6]. These activities were then classified into one of four activities intensities: sedentary, light, moderate, and vigorous. Walking activities (gardening, active commuting, etc) were classified by normalized gravitational units (g) where <100 milli g were classified as light intensity (<3 METs), ≥100 milli g and <400 milli g were considered moderate intensity physical activity (≥3 to <6 METs), and ≥400 milli g were considered vigorous-intensity PA (≥6 METs)[6]. All windows classified as running/high energetic activity were classified as vigorous-intensity physical activity (≥ 6 METs)[3,6,7]. Bouts of activity for moderate and vigorous intensity were classified as each event lasting 10 seconds or more.

### Sleep and non-wear time

The non-wear time was determined using a previously validated algorithm that uses wrist tilt angle to determine non-wear with 86-95% accuracy[4]. No values were imputed for non-wear time. Sleep was defined as the average daily duration of sleep (hours/day) as calculated using a validated algorithm based on relative changes in wrist tilt angle between successive 5-second windows[5]. For each interval of 5 seconds, the average of the estimated wrist tilt angle was calculated and a rolling 5-minute median served as an input for the algorithm to identify sleep onset and sleep offset, and then time spent asleep within this timeframe[4,5].

**Physical Activity Classification Scheme (Diagram A).**

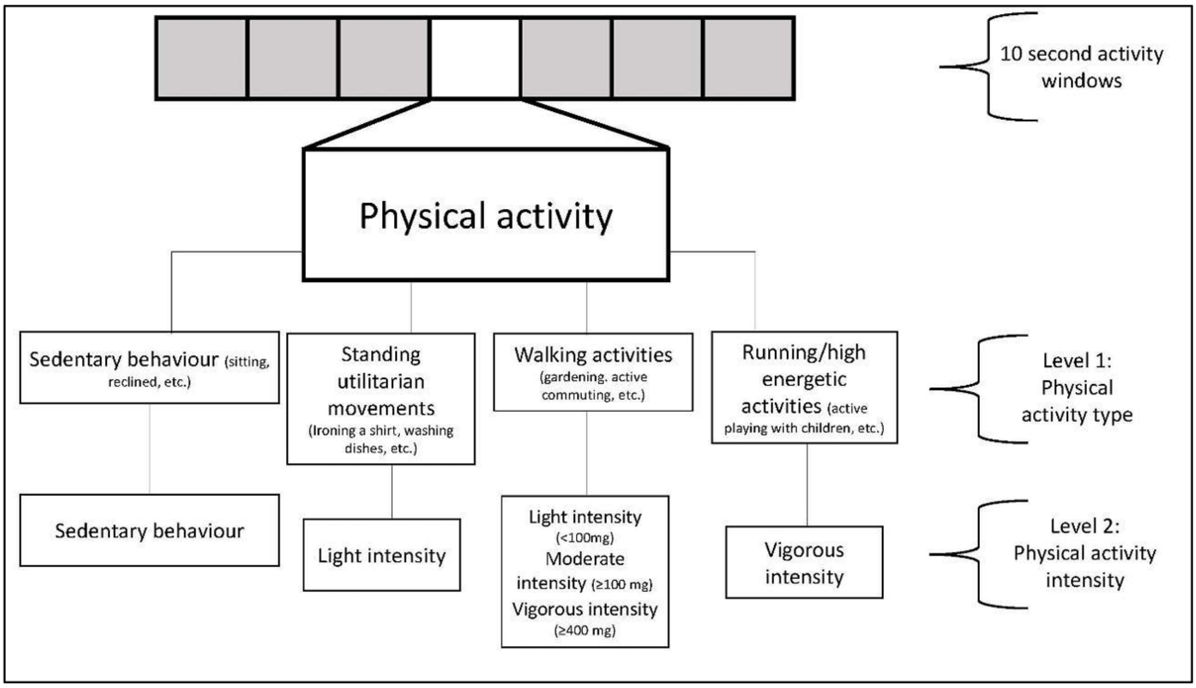


**Physical Activity Classification Performance**

The performance of this physical activity classification scheme was tested in an independent sample of 102 adults from the US[8] and Australia[9]. This data includes direct observation measurement of 105,767 activity samples from structured and free-living activities (17,627 minutes) which were used to test the robustness and generalizability of the two-stage activity and intensity classifier. The data was collected from participant-worn or researcher-held Go-Pro video recordings. All data was imported into Noldus Observer XT software for continuous video coding. The direct observation coding generated continuous physical activity codes corresponding to the start and finish of each movement. These coded movements were then compared against the accelerometer data using the available time-stamp information. The below table includes the performance metrics across activities. Interobserver reliability was assessed by dual coding. The intraclass correlation coefficient for coding activities was 0.912 (0.866-0.942). The performance in metrics and confusion matrix for activity classification is shown below.

**Classifier Performance Metrics for Intensity in US and Australian Adults**

| | Sensitivity | Specificity | Precision | F-score | Overall Accuracy | Weighted Kappa | Overall F-score |
|---|---|---|---|---|---|---|---|
| Sedentary | 86.5 | 93.7 | 90.5 | 88.5 | | | |
| Light | 71.2 | 89.4 | 55.8 | 62.6 | | | |
| Moderate | 85.4 | 96.6 | 92.7 | 88.9 | | | |
| Vigorous | 95.4 | 99.4 | 94.6 | 95.0 | | | |
| | | | | | **84.6** | 0.78 | 83.8 |

Rows= ground truth; columns=predictions; bold=correct classification; all activities were free-living or simulated free-living activities.

**Confusion Matrix for Activity Classification in US and Australian Adults**

| | Sedentary | Light | Moderate | Vigorous |
|---|---|---|---|---|
| Sedentary | **36,904** | 5,232 | 508 | 2 |
| Light | 3,120 | **11,712** | 1,612 | 17 |
| Moderate | 502 | 4,016 | **29,528** | 526 |
| Vigorous | 226 | 17 | 214 | **9,470** |

Rows= ground truth; columns=predictions; bold=correct classification; all activities were free-living or simulated free-living activities.

**Supplement Table 1**: Questions to determine leisure time physical activity

| Variable | Definition |
|---|---|
| Vigorous recreational activities (NHANES Survey ID: PAQ650) | The next questions exclude the work and transportation activities that you have already mentioned. Now I would like to ask you about sports, fitness and recreational activities. Do you do any vigorous-intensity sports, fitness, or recreational activities that cause large increases in breathing or heart rate like running or basketball for at least 10 minutes continuously? • Yes • No • Refused •Don't know |
| Days of vigorous recreational activities (NHANES Survey ID: PAQ655) | In a typical week, on how many days do you do vigorous-intensity sports, fitness or recreational activities? |
| Minutes vigorous recreational activities (NHANES Survey ID: PAQ660) | How much time do you spend doing vigorous-intensity sports, fitness or recreational activities on a typical day? |
| Moderate recreational activities (NHANES Survey ID: PAQ665) | Do you do any moderate-intensity sports, fitness, or recreational activities that cause a small increase in breathing or heart rate such as brisk walking, bicycling, swimming, or golf for at least 10 minutes continuously? • Yes • No • Refused •Don't know |
| Days of moderate recreational activities (NHANES Survey ID: PAQ670) | In a typical week, on how many days do you do moderate-intensity sports, fitness or recreational activities? |
| Minutes moderate recreational activities (NHANES Survey ID: PAQ675) | How much time do you spend doing moderate-intensity sports, fitness or recreational activities on a typical day? |

**Supplement Table 2**: Mortality definitions and ICD-10 Codes

| Mortality Type | Definition and ICD-10 Codes |
|---|---|
| All-Cause Mortality | **Diseases of heart** (I00-I09, I11, I13, I20-I51)<br><br>**Malignant neoplasms** (C00-C97)<br><br>**Chronic lower respiratory diseases** (J40-J47)<br><br>**Cerebrovascular diseases** (I60-I69)<br><br>**Alzheimer's disease** (G30)<br><br>**Diabetes mellitus** (E10-E14)<br><br>**Influenza and pneumonia** (J09-J18)<br><br>**Nephritis, nephrotic syndrome and nephrosis** (N00-N07, N17-N19, N25-N27)<br><br>**Accidents or unintentional injuries**<br>(V01-V99: Transport Accidents<br>W00-W19: Falls<br>W20-W49: Exposure to Inanimate Mechanical Forces<br>W50-W64: Exposure to Animate Mechanical Forces<br>W65-W74: Accidental Drowning and Submersion<br>W75-W84: Other Accidental Threats to Breathing<br>W85-W99: Exposure to Electric Current, Radiation, and Extreme Ambient Air Temperature and Pressure<br>X00-X99: Exposure to Forces of Nature<br>Y85-Y86: Sequelae of External Causes of Morbidity and Mortality)<br><br>**All other deaths (residual)**<br>This includes all ICD-10 codes not listed in the above categories[10] |
| Accidents or unintentional injuries | V01-V99: Transport Accidents<br>W00-W19: Falls<br>W20-W49: Exposure to Inanimate Mechanical Forces<br>W50-W64: Exposure to Animate Mechanical Forces<br>W65-W74: Accidental Drowning and Submersion |

| | |
|---|---|
| | W75-W84: Other Accidental Threats to Breathing<br>W85-W99: Exposure to Electric Current, Radiation, and Extreme Ambient Air Temperature and Pressure<br>X00-X99: Exposure to Forces of Nature<br>Y85-Y86: Sequelae of External Causes of Morbidity and Mortality |

ICD-10 codes were classified by the Data Linkage program into the following categories: Diseases of heart (I00-I09, I11, I13, I20-I51), Malignant neoplasms (C00-C97), Chronic lower respiratory diseases (J40-J47), Cerebrovascular diseases (I60-I69), Alzheimer's disease (G30), Diabetes mellitus (E10-E14), Influenza and pneumonia (J09-J18), Nephritis, nephrotic syndrome, and nephrosis (N00-N07, N17-N19, N25-N27), accidents or unintentional (V01-X59, Y85-Y86)[10,11]. The specific ICD-10 codes for events inside of each category are not publicly available.

**Supplement Table 3:** Covariate definitions

| Variable | Definition | NHANES Survey ID (if applicable) |
|---|---|---|
| Age | Continuous | RIDAGEYR |
| Sex | Female/Male | RIAGENDR |
| Ethnicity | Mexican American, Other Hispanic, White, Black, Asian, Other | RIDRETH3 |
| Education | Less than 9th grade, 9-11th grade (Includes 12th grade with no diploma), High school graduate/GED or equivalent, Some college or AA degree, College graduate or above | DMDEDUC2 |
| Household income | Household income to poverty ratio | INDFMPIR |
| Smoking status | Current, previous, never | SMQ020, SMQ040 |
| Alcohol consumption | Consumed at least 12 alcoholic beverages in the past year: Yes/No | ALQ101 |
| Light-intensity physical activity | Standing utilitarian movements, slow walking (<3 METs) | Derived from accelerometer data |
| Moderate-intensity physical activity | Brisk walking, energetic activities (≥3 to <6 METs) | Derived from accelerometer data |
| Sleep | Hours/day | Derived from accelerometer data |
| Fruit and vegetable intake | Fruit and vegetable intake servings/day as the average of two 24-hour dietary recalls | DR1TOT_G, DR2TOT_G |
| Healthy Eating Index 2015 | The Healthy Eat Index 2015[12] is a thirteen-component diet quality score including total fruit, whole fruit, vegetable, green bean, whole grain, dairy, protein foods, seafood and plant proteins, fatty acids, refined grains, sodium, added sugar, and saturated fat. Continuous; 0-100 (higher denotes higher diet quality) derived from the average of two 24-hour dietary recalls. | DR1TOT_G, DR2TOT_G |
| Use of cholesterol medication | Taking prescription for high cholesterol: Yes/No | BPQ080, BPQ100D |
| Use of blood pressure medication | Taking prescription for hypertension: Yes/No | BPQ020, BPQ040A |
| Use of diabetes medication | Taking insulin or other diabetic pills to lower blood sugar: Yes/No | DIQ050, DIQ070 |

| | | |
|---|---|---|
| Previous CVD | Ever told you have congestive heart failure, coronary heart disease, heart attack, or stroke: Yes/No | MCQ160B, MCQ160C, MCQ160D, MCQ160E, MCQ160F, |
| Previous cancer | Ever told you have a cancer or malignancy: Yes/No | MCQ220 |
| Previous diabetes | Ever told you have diabetes: Yes/No | DIQ010 |
| Self-reported health status | Excellent, Very good, Good, Fair, Poor | HSD010 |
| Familial history of cardiovascular disease | Self-reported close biological relative with cardiovascular disease: Yes/No | MCQ300A |
| Familial history of diabetes | Self-reported close biological relative with diabetes: Yes/No | MCQ300C |
| Body mass index | Continuous; kilogram/meter$^2$ | BMXBMI |
| Waist circumference | Continuous; cm | BMXWAIST |
| Fasting triglycerides | Continuous; mg/dL | LBXTR |
| Fasting total cholesterol | Continuous; mg/dL | LBXTC |
| Systolic blood pressure | Average of four measurements. Continuous; mmHg | BPXSY1, BPXSY2, BPXSY3, BPXSY4 |
| Diastolic blood pressure | Average of four measurements. Continuous; mmHg | BPXDI1, BPXDI2, BPXDI3, BPXDI4 |

**Supplement Table 4:** Model variance inflation factors

| VILPA Frequency Model | |
|---|---|
| **Variable** | **Variance inflation factor (VIF)** |
| VILPA Frequency (bouts lasting <1 minute) (accelerometry) | 2.55 |
| Vigorous energy expenditure (accelerometry) | 1.36 |
| Moderate physical activity energy expenditure (accelerometry) | 2.09 |
| Light physical activity energy expenditure (accelerometry) | 1.66 |
| Sleep (accelerometry) | 1.09 |
| Age (self-report) | 1.80 |
| Sex (self-report) | 1.35 |
| Ethnicity (self-report) | 1.61 |
| Smoking (self-report) | 1.60 |
| Diet (self-report) | 1.12 |
| Alcohol (self-report) | 1.41 |
| Education (self-report) | 1.61 |
| Discretionary screen time (self-report) | 1.15 |
| Previous CVD | 1.27 |
| Previous cancer | 1.16 |
| Previous type-two-diabetes | 1.45 |
| Familial history of CVD (self-report) | 1.06 |
| Familial history of cancer (self-report) | 1.23 |
| Medication (self-report) | 1.42 |
| **VILPA Duration Model** | |
| **Variable** | **Variance inflation factor (VIF)** |
| VILPA duration (bouts lasting <1 minute) (accelerometry) | 2.66 |
| VILPA duration (bouts lasting>1 minute) (accelerometry) | 1.55 |
| Moderate physical activity energy expenditure (accelerometry) | 2.07 |
| Light physical activity energy expenditure (accelerometry) | 1.63 |
| Sleep (accelerometry) | 1.09 |
| Age (self-report) | 1.77 |
| Sex (self-report) | 1.35 |
| Ethnicity (self-report) | 1.63 |
| Smoking (self-report) | 1.60 |
| Diet (self-report) | 1.13 |
| Alcohol (self-report) | 1.42 |
| Education (self-report) | 1.62 |
| Discretionary screen time (self-report) | 1.16 |
| Previous CVD | 1.28 |
| Previous cancer | 1.16 |
| Previous type-two-diabetes | 1.45 |
| Familial history of CVD (self-report) | 1.06 |
| Familial history of cancer (self-report) | 1.16 |
| Medication (self-report) | 1.41 |

The table provides the variance inflation factor (VIF) for each covariate in the primary analytical model. VIF values measure multicollinearity among the exposure variables, with a value of 1 indicating no correlation with other exposures. Higher values suggest increasing multicollinearity, with values greater than 5 indicating potentially problematic multicollinearity[1]. Cardiovascular disease (CVD); Vigorous physical activity (VPA); Vigorous intermittent lifestyle physical activity (VILPA).

**Supplement Table 5**: Interaction test between sex and VILPA with all-cause mortality

| Sex*VILPA Model | | | |
|---|---|---|---|
| **VILPA Frequency** | | | |
| **Outcome** | **Multiplicative interaction** | | **Additive relative excess risk due to interaction (RERI)** |
| | HR (95% CI) | p-value | RERI % (95%CI) |
| All-Cause Mortality | 0.93 (0.81, 1.07) | 0.30 | -0.01 (-0.49, 0.46) |
| **VILPA Duration** | | | |
| **Outcome** | **Multiplicative interaction** | | **Additive relative excess risk due to interaction (RERI)** |
| | HR (95% CI) | p-value | RERI % (95%CI) |
| All-Cause Mortality | 1.48 (0.77, 2.82) | 0.24 | -0.06 (-1.18, 1.04) |

The table shows the hazard ratios for the multiplication interaction term (e.g., VILPA daily duration*sex) in the model and the relative excess risk due to interaction (RERI) for the frequency and total daily duration of VILPA[13]. The models were adjusted for sex, age, income, education, ethnicity, fruit and vegetable consumption, smoking history, physical activity energy expenditure from LPA and MPA, alcohol consumption, sleep duration, PAEE from light and moderate intensity, discretionary screentime, medication use (glycaemic control, blood pressure, cholesterol), family history of diabetes and CVD, and previous history of CVD, diabetes, and cancer. The VILPA frequency model was additionally adjusted for PAEE from vigorous-intensity physical activity and the VILPA daily duration model was additionally adjusted for VPA duration from bouts lasting >1 minute in duration.

**Supplement Table 6.** E-values for VILPA frequency and duration

A. VILPA Frequency

| VILPA Frequency | E-Value |
|---|---|
| All-cause Mortality | 3.59 (1.96) |

B. VPA Duration

| VILPA Duration | E-Value |
|---|---|
| All-cause Mortality | 3.59 (2.00) |

Values represent the point estimate and lower limit of the confidence interval (value is equal to 1 if crossing the referent line) in brackets that an unmeasured confounder would need to have with both the exposure and outcome, conditional on the measured covariates to explain away the exposure-outcome association[14]. Vigorous intermittent lifestyle physical activity (VILPA).

**Supplement Table 7:** STROBE statement

| | Item No | Recommendation | Page No |
|---|---|---|---|
| **Title and abstract** | 1 | (a) Indicate the study's design with a commonly used term in the title or the abstract | 1-3 |
| | | (b) Provide in the abstract an informative and balanced summary of what was done and what was found | 1-3 |
| **Introduction** | | | |
| Background/rationale | 2 | Explain the scientific background and rationale for the investigation being reported | 5-6 |
| Objectives | 3 | State specific objectives, including any prespecified hypotheses | 6 |
| **Methods** | | | |
| Study design | 4 | Present key elements of study design early in the paper | 7-13 |
| Setting | 5 | Describe the setting, locations, and relevant dates, including periods of recruitment, exposure, follow-up, and data collection | 7 |
| Participants | 6 | (a) Give the eligibility criteria, and the sources and methods of selection of participants. Describe methods of follow-up | 7-8 |
| | | (b) For matched studies, give matching criteria and number of exposed and unexposed | NA |
| Variables | 7 | Clearly define all outcomes, exposures, predictors, potential confounders, and effect modifiers. Give diagnostic criteria, if applicable | 8-9 |
| Data sources/ measurement | 8* | For each variable of interest, give sources of data and details of methods of assessment (measurement). Describe comparability of assessment methods if there is more than one group | 7-9 |
| Bias | 9 | Describe any efforts to address potential sources of bias | 11 |

| | | | |
|---|---|---|---|
| Study size | 10 | Explain how the study size was arrived at | Supplementary figure 1 |
| Quantitative variables | 11 | Explain how quantitative variables were handled in the analyses. If applicable, describe which groupings were chosen and why | 9 |
| Statistical methods | 12 | (a) Describe all statistical methods, including those used to control for confounding | 10-13 |
| | | (b) Describe any methods used to examine subgroups and interactions | 12-13 |
| | | (c) Explain how missing data were addressed | 12 |
| | | (d) If applicable, explain how loss to follow-up was addressed | 12-13 |
| | | (e) Describe any sensitivity analyses | 13 |
| **Results** | | | |
| Participants | 13* | (a) Report numbers of individuals at each stage of study—eg numbers potentially eligible, examined for eligibility, confirmed eligible, included in the study, completing follow-up, and analysed | 13 |
| | | (b) Give reasons for non-participation at each stage | Supplemental figure 1 |
| | | (c) Consider use of a flow diagram | Supplemental figure 1 |
| Descriptive data | 14* | (a) Give characteristics of study participants (eg demographic, clinical, social) and information on exposures and potential confounders | Table 1 |
| | | (b) Indicate number of participants with missing data for each variable of interest | Supplemental figure 1 |
| | | (c) Summarise follow-up time (eg, average and total amount) | Table 1 |
| Outcome data | 15* | Report numbers of outcome events or summary measures over time | Supplemental figure 1<br>Table 1 |

| | | | |
|---|---|---|---|
| Main results | 16 | (a) Give unadjusted estimates and, if applicable, confounder-adjusted estimates and their precision (eg, 95% confidence interval). Make clear which confounders were adjusted for and why they were included | 14-15 |
| | | (b) Report category boundaries when continuous variables were categorized | NA |
| | | (c) If relevant, consider translating estimates of relative risk into absolute risk for a meaningful time period | 14 |
| Other analyses | 17 | Report other analyses done—eg analyses of subgroups and interactions, and sensitivity analyses | 16-17 |
| **Discussion** | | | |
| Key results | 18 | Summarise key results with reference to study objectives | 17-18 |
| Limitations | 19 | Discuss limitations of the study, taking into account sources of potential bias or imprecision. Discuss both direction and magnitude of any potential bias | 21-23 |
| Interpretation | 20 | Give a cautious overall interpretation of results considering objectives, limitations, multiplicity of analyses, results from similar studies, and other relevant evidence | 23 |
| Generalisability | 21 | Discuss the generalisability (external validity) of the study results | 22-23 |
| **Other information** | | | |
| Funding | 22 | Give the source of funding and the role of the funders for the present study and, if applicable, for the original study on which the present article is based | 24 |

*Give information separately for exposed and unexposed groups.

**Note:** An Explanation and Elaboration article discusses each checklist item and gives methodological background and published examples of transparent reporting. The STROBE checklist is best used in conjunction with this article (freely available on the Web sites of PLoS Medicine at http://www.plosmedicine.org/, Annals of Internal Medicine at http://www.annals.org/, and Epidemiology at http://www.epidem.com/). Information on the STROBE Initiative is available at http://www.strobe-statement.org.

### References

1. Ahmadi MN, Holtermann A, Tudor-Locke C, et al. Time to Elicit Physiological and Exertional Vigorous Responses from Daily Living Activities: Setting Foundations of an Empirical Definition of VILPA. Med Sci Sports Exerc. 2024;doi:10.1249/mss.0000000000003521
2. Stamatakis E, Ahmadi MN, Friedenreich CM, et al. Vigorous Intermittent Lifestyle Physical Activity and Cancer Incidence Among Nonexercising Adults: The UK Biobank Accelerometry Study. JAMA Oncology. 2023;9(9):1255-1259. doi:10.1001/jamaoncol.2023.1830
3. Stamatakis E, Ahmadi MN, Gill JMR, et al. Association of wearable device-measured vigorous intermittent lifestyle physical activity with mortality. Nature Medicine. 2022;28(12):2521-2529. doi:10.1038/s41591-022-02100-x
4. Ahmadi MN, Nathan N, Sutherland R, Wolfenden L, Trost SG. Non-wear or sleep? Evaluation of five non-wear detection algorithms for raw accelerometer data. Journal of Sports Sciences. 2020;38(4):399-404.
5. van Hees VT, Sabia S, Jones SE, et al. Estimating sleep parameters using an accelerometer without sleep diary. Scientific Reports. 2018;8(1):12975. doi:10.1038/s41598-018-31266-z
6. Pavey TG, Gilson ND, Gomersall SR, Clark B, Trost SG. Field evaluation of a random forest activity classifier for wrist-worn accelerometer data. Journal of Science and Medicine in Sport. 2017;20(1):75-80. doi:10.1016/j.jsams.2016.06.003
7. Ahmadi MN, Hamer M, Gill JM, et al. Brief bouts of device-measured intermittent lifestyle physical activity and its association with major adverse cardiovascular events and mortality in people who do not exercise: a prospective cohort study. The Lancet Public Health. 2023;8(10):e800-e810.
8. Reiss A, Weber M, Stricker D. Exploring and extending the boundaries of physical activity recognition. 2011:46-50.
9. Clark BK, Winkler EA, Brakenridge CL, Trost SG, Healy GN. Using Bluetooth proximity sensing to determine where office workers spend time at work. PLOS ONE. 2018;13(3):e0193971. doi:10.1371/journal.pone.0193971
10. Anderson RN, Miniño AM, Hoyert DL, Rosenberg HM. Comparability of cause of death between ICD-9 and ICD-10: preliminary estimates. Natl Vital Stat Rep. May 18 2001;49(2):1-32.
11. National Center for Health Statistics Division of Analysis and Epidemiology. Continuous NHANES Public-use Linked Mortality Files, 2019. https://www.cdc.gov/nchs/data-linkage/mortality-public.htm
12. Krebs-Smith SM, Pannucci TE, Subar AF, et al. Update of the Healthy Eating Index: HEI-2015. J Acad Nutr Diet. Sep 2018;118(9):1591-1602. doi:10.1016/j.jand.2018.05.021
13. Andersson T, Alfredsson L, Källberg H, Zdravkovic S, Ahlbom A. Calculating measures of biological interaction. European Journal of Epidemiology. 2005;20(7):575-9. doi:10.1007/s10654-005-7835-x
14. Haneuse S, VanderWeele TJ, Arterburn D. Using the E-Value to Assess the Potential Effect of Unmeasured Confounding in Observational Studies. JAMA. 2019;321(6):602-603. doi:10.1001/jama.2018.21554
